# Combined GWAS and single cell transcriptomics uncover the underlying genes and cell types in disorders of gut-brain interaction

**DOI:** 10.1101/2023.06.02.23290906

**Authors:** Alireza Majd, Mikayla N Richter, Ryan M Samuel, Andrius Cesiulis, Zaniar Ghazizadeh, Jeffrey Wang, Faranak Fattahi

## Abstract

Disorders of gut-brain interaction (DGBIs), formerly known as functional gastrointestinal disorders, are extremely common and historically difficult to manage. This is largely because their cellular and molecular mechanisms have remained poorly understood and understudied. One approach to unravel the molecular underpinnings of complex disorders such as DGBIs is performing genome wide association studies (GWASs). However, due to the heterogenous and non-specific nature of GI symptoms, it has been difficult to accurately classify cases and controls. Thus, to perform reliable studies, we need to access large patient populations which has been difficult to date. Here, we leveraged the UK Biobank (UKBB) database, containing genetic and medical record data of over half a million individuals, to perform GWAS for five DGBI categories: functional chest pain, functional diarrhea, functional dyspepsia, functional dysphagia, and functional fecal incontinence. By applying strict inclusion and exclusion criteria, we resolved patient populations and identified genes significantly associated with each condition. Leveraging multiple human single-cell RNA-sequencing datasets, we found that the disease associated genes were highly expressed in enteric neurons, which innervate and control GI functions. Further expression and association testing-based analyses revealed specific enteric neuron subtypes consistently linked with each DGBI. Furthermore, protein-protein interaction analysis of each of the disease associated genes revealed protein networks specific to each DGBI, including hedgehog signaling for functional chest pain and neuronal function and neurotransmission for functional diarrhea and functional dyspepsia. Finally, through retrospective medical record analysis we found that drugs that inhibit these networks are associated with an increased disease risk, including serine/threonine kinase 32B drugs for functional chest pain, solute carrier organic anion transporter family member 4C1, mitogen-activated protein kinase 6, and dual serine/threonine and tyrosine protein kinase drugs for functional dyspepsia, and serotonin transporter drugs for functional diarrhea. This study presents a robust strategy for uncovering the tissues, cell types, and genes involved in DGBIs, presenting novel predictions of the mechanisms underlying these historically intractable and poorly understood diseases.

## Introduction

Disorders of gut-brain interactions (DGBIs) formerly known as functional gastrointestinal (GI) disorders, consist of a wide spectrum of conditions, including disorders that affect the upper GI (e.g. functional dyspepsia, functional heartburn, functional dysphagia) or lower GI (e.g. irritable bowel syndrome (IBS), functional constipation, functional fecal incontinence). DGBIs are the most common diagnoses in gastroenterology^1^. According to recent studies, 37-40% of participants worldwide meet the diagnostic criteria for one or more DGBI^2, 3^. DGBIs are chronic diseases characterized by symptoms such as pain, bloating, constipation, diarrhea, and nausea^4, 5^. They can significantly reduce quality of life due to the severity of these symptoms, and current treatment options have limited efficacy^6^. Patients with DGBI often resort to pain medications to alleviate their symptoms, and they tend to use prescription analgesics twice as frequently^4^. Unfortunately, this increased medication use can result in other GI side effects, including opioid-induced constipation^5^.

DGBI symptoms are complex, largely non-specific, chronic, and progressive, making them difficult to diagnose efficiently and effectively. In addition, patients’ subjective thresholds for using negative affective terms when describing GI discomfort make diagnosis even more challenging^6, 7^. Furthermore, similar symptoms could be attributed to other GI disorders such as the esophageal webs, IBD, lactose intolerance, GI malignancies, malabsorption syndromes, food allergies, GI infections, peptic ulcer disease, postsurgical complications, biliary pain, drug-induced side effects, and so on. It is also possible for non-GI diseases to mimic DGBIs, for example, chronic abdominal pain may be caused by endometriosis, and difficulty in swallowing and constipation may be early symptoms of Parkinson’s disease. Hence, extensive diagnostic investigations are usually performed, including upper and lower endoscopies, in order to exclude more serious conditions that show similar symptoms^8^.

DGBI associated GI motility disturbances can be caused by visceral hypersensitivity and changes in mucosal immunity, gut microbiota, and the central nervous system^9, 10^. However, the general consensus is that abnormalities in the enteric nervous system (ENS) are responsible for many DGBIs^11, 12^. The ENS is the largest division of the peripheral nervous system that is embedded in the GI tract and control its functions. It is composed of a diverse array of enteric neurons and glia that regulate a myriad of activities to maintain gut homeostasis, including motility, secretion, and immune response^13^. However, due to the inaccessibility of the ENS, which makes up approximately 1% of GI tissue^14^, mechanistic studies to uncover the molecular and cellular pathophysiology of DGBIs has been technically infeasible. Nevertheless, previous studies have identified changes in the ENS in specific forms of DGBIs, such as IBS and functional dyspepsia. In IBS, there is an increase in density and sprouting of nerve fibers in mucosal tissues and abdominal pain is positively correlated with the number of mast cells close to enteric neurons^15, 16^. In functional dyspepsia, the submucosal plexus within the GI is abnormal, with eosinophils and mast cell infiltration occurring at a higher rate than controls^17^. However, due to the limitations of DGBI animal models^10, 18–20^ and challenges in obtaining patient tissue samples in large enough numbers for mechanistic studies, progress toward defining the cell types and protein pathways responsible for DGBI pathophysiology has been slow. Furthermore, due to the complexity of nerve-tissue interactions studied *in vivo*, it remains unclear which GI cell types and intracellular mechanisms drive disease pathophysiology.

Recently, multiple single cell transcriptomics datasets of the human GI tract^14, 21^ and the ENS^22, 23^ have been published, enabling the prediction of cell types associated with DGBIs based on their unique transcriptional features. However, the genetic underpinnings of DGBIs have remained elusive, making the integrating of transcriptomic data with DGBI studies unfeasible to date.

Recently, there have been efforts to shed light on the genetics of these disorders by performing genome wide association study (GWAS), but other than IBS^24–28^, other DGBI categories have received less attention and causal genetic variants have yet to be identified^29^. This is likely because many DGBIs are difficult to accurately diagnose, which increases the chance of mislabeling in retrospective datasets, making it challenging to accurately define cases and controls. There has been increasing precedence supporting the use of symptom-based diagnosis as opposed to diagnosing solely when organic diseases have been excluded^12^. The Rome IV criteria^9^ therefore require less strict exclusions by replacing “no evidence of organic disease” with “symptoms cannot be attributed to another medical condition”^30–32^. Nonetheless, in order to accurately identify cases and controls for complex diseases such as DGBIs, large cohorts are an important requirement^33^.

Here, we leveraged the large-scale UK Biobank (UKBB) biomedical database, which contains genetic and health information from over half a million patients in the United Kingdom, to perform GWASs on five DGBIs. These include functional chest pain, functional diarrhea, functional dyspepsia, functional dysphagia, and functional fecal incontinence. By applying extensive exclusion criteria to remove potential overlaps and decrease noise, we identified single nucleotide polymorphisms (SNPs) and their implicated genes significantly associated with each DGBI. In order to enhance accessibility of these results for the research community, we developed an online tool on BBNbrowser.com. This tool enables researchers to explore and determine the DGBI disease association for any gene of interest. Next, by assessing expression patterns in human GI transcriptomic datasets, we determined the tissues and cell types associated with each DGBI, linking enteric neurons and specific neuronal subtypes with each DGBI. Furthermore, by mapping the interactions of the DGBI associated proteins expressed in the implicated neurons, we uncovered DGBI associated cellular pathways. Lastly, we leveraged electronic medical record data from the UCSF De-Identified Clinical Data Warehouse^34^, to evaluate the clinical relevance of these pathways and their potential therapeutic significance. Remarkably, patients that received drugs that antagonize or inhibit these pathways were significantly more likely to have a DGBI diagnosis, validating the link between these pathways and DGBI pathophysiology. Altogether, this study provides a framework for identifying the specific tissues, cell types, and proteins affected in DGBIs, which may improve disease management and enable the design of new and effective therapies for these historically intractable conditions.

## Results

### Defining DGBI study groups in the UKBB based on Rome IV criteria

Performing case-control GWAS of DGBIs is challenging due to frequent misdiagnoses. Therefore, studies with small sample sizes or non-stringent case-control definitions may have limited power. In our study, we aimed to overcome this challenge by leveraging the large-scale UKBB database, excluding patients with conditions known to cause similar symptoms to the DGBI of interest, thereby enhancing the resolution for defining cases and controls (Figure 1A). All the corresponding conditions were filtered based on primary care and hospital inpatient data, self-report diseases, cancer registries and history of surgeries which can cause complications mimicking the DGBIs (Table S1). UKBB participants (n = 28,052) met the criteria for at least one of the five studied DGBIs. Functional chest pain showed the highest prevalence (4.5%) among participants remaining in cohort after disease-specific exclusions, followed by functional dyspepsia (1.7%), functional dysphagia (1.1%), functional fecal incontinence (0.3%), and functional diarrhea (<0.1%). After sample quality controls, 19,776 cases remained with at least one DGBI in their records for genetic association analysis (Figure 1B). In comparison with previous community based reports based on Rome IV criteria^5^, this cohort has lower prevalence of most disorders, except functional chest pain, which is 3.1% more prevalent. Functional dyspepsia, functional dysphagia, functional fecal incontinence and functional diarrhea had lower prevalence by 5.5%, 2.1%, 1.3% and 4.6%, respectively. This reduction is likely due to our exclusion criteria for cases and controls that removed patients with other overlapping conditions^2^ .Male participants had a higher average age compared to female participants (p value <0.001) (Table S2). Female to male odds ratios (ORs) were significant in functional dysphagia OR= 1.32 (95% confidence interval: 1.22, 1.41), functional dyspepsia OR= 1.61 (95% confidence interval: 1.52, 1.72), functional fecal incontinence OR= 1.79 (95% confidence interval: 1.59, 2.04), and functional diarrhea OR= 1.85 (95% confidence interval: 1.05, 3.45), but not for functional chest pain OR= 0.99 (95% confidence interval: 0.95, 1.03) (Figure 1C).

**Figure 1:**
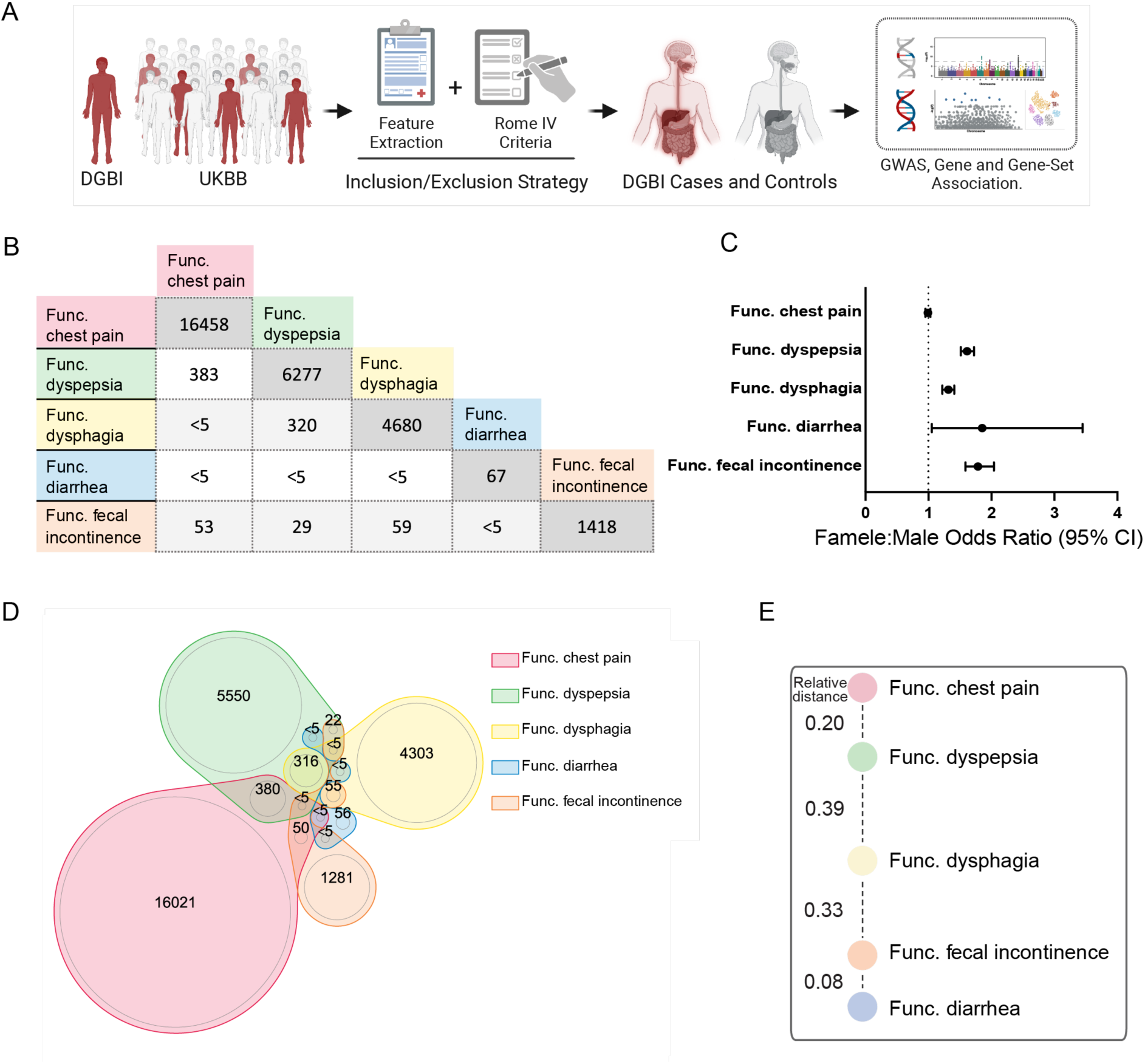
Cohort characteristics of functional chest pain, functional diarrhea, functional dyspepsia, functional dysphagia, and functional fecal incontinence in the UK Biobank. **A)** Schematic of the UKBB GWAS pipeline. **B)** An overview of cases shared by different disease groups. **C)** Female to male odds ratios across disease case groups and each corresponding control group. **D)** Venn diagram illustrating the distribution of cases among different groups. **E)** Comorbidity analysis results showing the relative distance between disease groups.

To evaluate the association between different disorders, we applied a sequential approach. We took one disorder as an index group, then compared the proportion of individuals within the remaining four disorder groups who were diagnosed with both the index and the second disorder. To determine which disorder was most likely to be comorbid with each index disorder, we used the two-proportion Z test to compare the fraction of shared cases among all comparison groups (Figure S1). The association z-scores highlighted an anatomical basis for comorbidities, where the patients are more likely to suffer from multiple upper GI or lower GI disorders simultaneously (Figure 1E). A higher comorbidity score across disorders in closer anatomical regions is consistent with previous studies, highlighting the possibility of region-specific pathophysiological mechanisms that contribute to these diseases^35^. Overall, it is noteworthy that there is a relatively considerable number of cases with comorbidities in other DGBI categories (Figure 1D, Figure S1). To uncover the complexity of these disorders, it is crucial to understand that DGBIs do not manifest as independent conditions and are largely Interrelated. As a result, there is a need for diagnosis and treatment guidelines for individuals with more than one DGBI^6, 36^.

### Identification of genetic variants and genes associated with DGBIs

To identify SNPs and genes associated with each of the five DGBIs, we performed GWAS using Regenie^37^, including a sixth category representing a combined cohort (all DGBI) (Figure S2). We identified variants with *P*<5×10^-8^ for the functional fecal incontinence and the “all DGBI” GWAS, including intronic variant rs975577 located in *MRM1* gene and intergenic variant rs2081046 21K upstream to ENSG00000270212 (CTD-2540B15.12) transcript for functional fecal incontinence and intergenic vartiant rs114969851 on chromosome 5 with 18K distance to ARL2BPP6 gene for “all DGBI” (Figure S2, Figure 2A). For functional dyspepsia, functional chest pain, and functional diarrhea, the signals with the highest association level were intergenic variant rs60111579 on chromosome 5, intronic variant rs76952347 in LYPD6 gene on chromosome 2, and intergenic variant rs185001252 on chromosome 5 respectively (Figure S2, Figure 2A). Structural and functional annotations for all the variants with p < 10−6 are included in Table S3. In total, there were 162 genes that were mapped to potential variants and were used in downstream analyses (Table S3).

**Figure 2:**
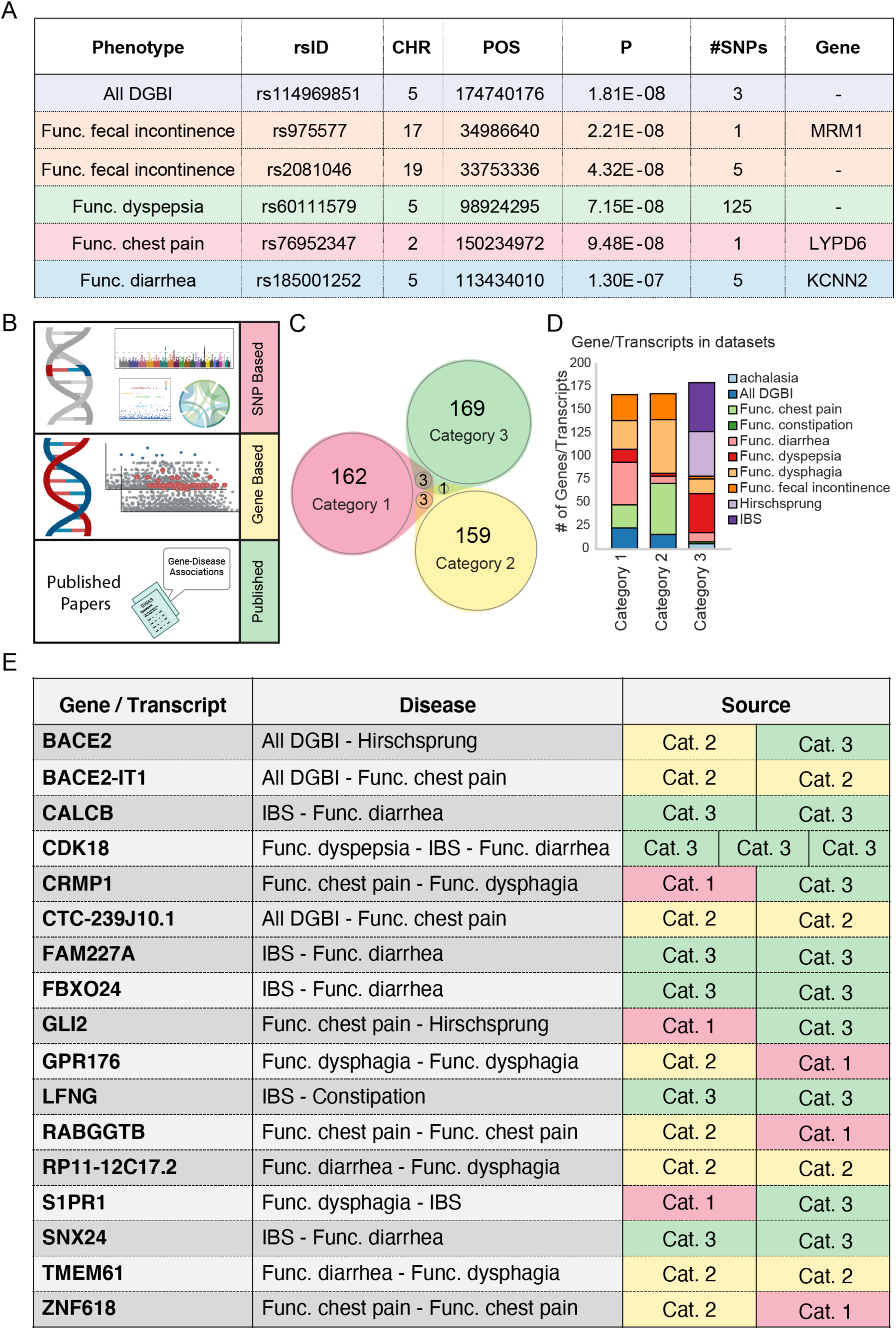
Genes associated with DGBIs map to relevant tissues and functional pathways in the GI. **A)** Top DGBI variants identified by GWAS and their closest gene. **B)** Schematic illustration of three categories of DGBI associated gene lists. 1. Genes mapped to SNP hits in our GWAS. 2. Genes filtered in via z-score threshold from gene-based association testing and 3. previously published GWAS. **C)** Venn diagram illustrating the overlap between DGBI associated gene categories. **D)** Stacked bar plot representing the composition of DGBI associated genes in each category. **E)** List of genes/transcripts in at least two out of three DGBI associated gene categories. Cat. 1: mapped genes to significant SNPs; Cat. 2: filtered from gene-based association testing; Cat. 3: from previously published GWAS.

For each disorder, we evaluated the enrichment of our mapped genes in 30 general tissue types using GTEx v8 30 data^38^ (Figure S3A). Functional chest pain-mapped genes were enriched in nerve tissue (p-adj = 0.046) (Figure S3A, B). In other disorders, no tissues were significantly enriched for mapped genes. Although, among the list of genes in functional dysphagia we observed high expression of *EPHA7* and *NGB* in the esophagus, colon, and stomach, and high expression of functional dyspepsia genes: *RGMB* in the stomach and esophagus, and *TRIM36* in small intestine, colon and stomach (Figure S3A). Interestingly, *EPHA7* and *TRIM36* have also been found to be associated with gut microbiota alterations in previous GWAS studies, highlighting the need for further research into the role of these genes in the crosstalk between microbiota and DGBIs^39, 40^.

We next leveraged the SNP-based data to conduct gene-based association testing and estimate the combined association of variants (including rare variants) located in one gene/transcript region. Using MAGMA^41^ we calculated the z-score values for a given gene as a measure of its disease association in the gene-level analyses. 159 genes with the strongest associations were filtered by z-score ± 3.5 (Table S4).

We next built a disease-associated list of genes for each DGBI by combining the genes annotated to significant SNPs (category 1), z-score filtered genes/transcripts from gene-based testing (category 2), and previously published GWAS results for related GI disorders^24, 25, 27–29, 42–58^ (category 3)(Figure 2B-D). The published category contained an expanded list of other potentially related disease categories, including achalasia, constipation, Hirschsprung’s disease and IBS (Figure 2D). Notably, these different approaches identified DGBI molecular features that were largely non-overlapping (Figure 2C), suggesting that combining multiple approaches is warranted to uncover different features of disease, particularly complex and poorly understood disorders like DGBIs. Genes identified in at least two of the three categories are listed in Figure 2E. Both SNP-based and gene-based analyses identified *GPR176* as a prominent gene for functional dysphagia, similar to *RABGGTB* and *ZNF618* for functional chest pain. Associations between *GPR176* and gut related phenotypes such as alterations in gut microbiome composition and diverticular disease have been reported previously^59, 60^. Moreover, *CDK18* has been previously linked to functional dyspepsia, IBS, and functional diarrhea, highlighting its relevance to DGBIs (Figure 2E).

### DGBI-associated genes are highly expressed in enteric neurons

To determine the molecular and cellular pathways relevant to each DGBI, we performed pre-ranked gene set enrichment analysis on the transcripts sorted based on z-scores (category 2) for each disorder. Based on a p-value cutoff of < 0.01, all five DGBI gene signatures showed enrichment for several pathways related to nervous system functions including neurotransmission-related gene sets, heavily implicating the ENS in the pathogenesis of these DGBIs (Figure 3A). Functional chest pain, functional diarrhea, and functional dyspepsia also had gene sets relating to muscle cells suggesting smooth muscle cells in the GI tract may be involved in the pathophysiology of these DGBIs (Figure 3A). Furthermore, this analysis identified gene sets relevant to multiple GI disorders, including intestinal hypoplasia for functional diarrhea, abnormal esophagus morphology, peptic ulcer, and tracheoesophageal fistula for functional dyspepsia, and dysphagia for functional fecal incontinence (Figure 3A). It is encouraging that pre-ranked gene set enrichment analysis of our GWAS-identified DGBI-associated genes found pathways relevant to the GI tract, suggesting that this unbiased approach can identify disease relevant cells and tissues. Additional pathways related to metabolism, including mitochondrial functions and lipid metabolism were significantly associated with functional chest pain, functional diarrhea, functional dyspepsia, and functional fecal incontinence (Figure S3C). Furthermore, pathways related to immune response were significantly enriched in all five DGBI categories, supporting the role of inflammation and immune system activity in DGBIs (Figure S3C). This is an intriguing observation since infections or auto-immune-induced inflammation are known to affect the ENS and cause abnormal GI motility, epithelial permeability), and enteric neuron damage and death, further supporting a mechanistic connection between the immune system, the ENS and DGBIs^13^.

**Figure 3:**
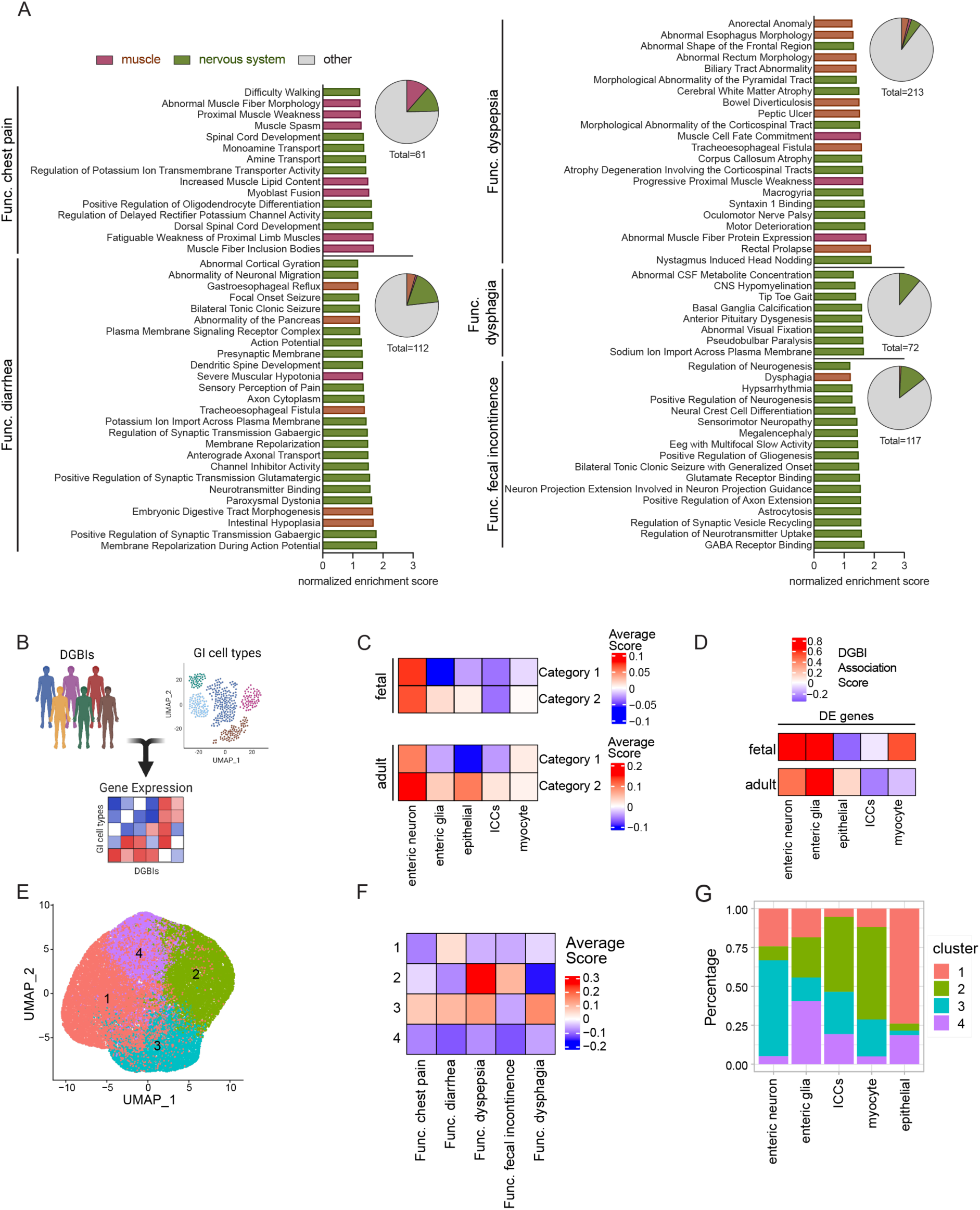
DGBI associated transcripts are enriched in enteric neurons. **A)** Summary of gene sets significantly enriched in the DGBI associated genes. The bar graphs highlight nervous system, muscle, and GI related pathways and the pie charts show the proportion of significantly enriched gene sets relating to these categories. **B)** Schematic illustration of the analysis design to evaluate expression of DGBI molecular features in GI cell types. **C)** Heatmaps of the average module scores of DGBI associated molecular features in human fetal (top) and adult (bottom) GI cell types using significantly associated transcripts from gene-based testing (z-score ± 3.5) and significantly associated genes from SNP-based testing (mapped genes). **D)** Heatmap of the gene-set enrichment analysis of DGBI associated genes among differentially expressed genes between clusters of human fetal and adult GI cell types. The values are calculated as the average of association z-scores for each cluster. **E)** UMAPs of the human fetal and adult GI cell types re-clustered based on their DGBI module scores. **F)** Heatmap of the average module scores of DGBI associated genes in the human fetal and adult GI cell types re-clustered based on their DGBI module scores. **G)** Stacked bar plot of the representation of human fetal and adult GI cell types in the clusters of human fetal and adult GI cell types re-clustered based on their DGBI module scores.

The GI tract is a complex organ system containing diverse cell types organized in multiple layers that all contribute to gut homeostasis. The mucosal layer is closest to the lumen and is primarily composed of epithelial cells, the longitudinal and circular muscle layers are primarily composed of myocytes and interstitial cells of cajal (ICCs), and the submucosal and myenteric plexuses are primarily composed of enteric neurons and glia^61^. Genetic abnormalities in any of these tissues could disrupt gut functions and potentially contribute to motility defects. To evaluate the expression of our DGBI-associated molecular features in these relevant cell types, we leveraged single cell transcriptomic data from human tissue samples (Figure 3B). We focused our analysis on enteric neurons, enteric glia, epithelial cells, ICCs, and myocytes from the single cell RNA sequencing (scRNA-seq) dataset of the primary fetal human intestine previously published by Teichmann and colleagues as well as the single nuclei RNA sequencing (snRNA-seq) dataset of the primary adult human colon previously published by Drokhlyansky et al. and Elmentaite et al. ^14, 21^(Figure S4A).

To predict the specific cell types affected by DGBIs, we utilized our curated DGBI transcript lists, including the transcripts resulting from our own GWAS (categories 1 and 2) and published GWAS analyses (category 3) (Table S5). We then module scored the cell types based on their expression of the molecular features associated with each DGBI. Interestingly, both category 1 and 2 transcripts showed the highest expression in enteric neurons in both datasets (Figure 3C). For the fetal dataset, this was primarily driven by a high expression of transcripts associated with functional chest pain, functional diarrhea, functional dyspepsia, and functional dysphagia (Figure S4B). For the adult dataset, the high module score for enteric neurons was driven by category 1 transcripts associated with functional diarrhea, functional fecal incontinence, and functional dysphagia and category 2 transcripts associated with functional chest pain, functional diarrhea, functional dyspepsia, and functional dysphagia (Figure S4C). Notably, functional fecal incontinence only had a positive module score for enteric neurons when the transcripts were evaluated in the adult dataset, in all other evaluations the module score was negative, suggesting that this DGBI may be affected by multiple cell types, including epithelial cells and myocytes which scored positively for its associated transcripts (Figure S4B and Figure S4C). To validate our observations, we utilized our curated list of category 3 transcripts identified through independently published GWAS studies^24,25,27–29,42–58(Table^ S5). Module scoring the GI cell types based on their expression of this more diverse set of DGBI phenotypes, we again observed that enteric neurons have a positive overall module score, although not the highest as we saw reflected by our own analyses (Figure S4D-E). Across both the fetal and adult datasets, functional dyspepsia consistently scored highly for enteric neurons, replicating the results from the category 1 and 2 analyses (Figure S4B-E). Additionally, similar to our previous analysis, functional fecal incontinence consistently scored negatively (Figure S4B-E). It is remarkable that DGBI module scores from the different transcript lists highlight the same cell types given that very few individual transcripts are shared among the different categories (Figure 2C). Thus, the score is not driven by an overlap between the transcripts identified in each category but instead points to broad biological features that implicate these cell types.

For the additional DGBI associated genes published previously^24, 25, 27–29, 42–58^, we observed that achalasia, constipation, and IBS scored negatively in the fetal dataset but showed a positive score in the adult dataset for enteric neurons (Figure S4D-E). This could suggest an age-dependent molecular shift occurring in enteric neurons, making them resistant to developing these DGBIs early in life and more susceptible to developing them in adulthood. Notably, achalasia, constipation, and IBS are more commonly diagnosed in adults rather than children^62–64^.

Altogether, our module score-based analyses implicate enteric neurons in DGBIs, most specifically functional chest pain, functional diarrhea, functional dyspepsia, and functional dysphagia and less specifically functional fecal incontinence. To further evaluate these findings through an alternative approach, we leveraged each cell-type’s differentially expressed genes from each dataset. Using the effect sizes of gene-level associations, we calculated the correlation of different GI cell types with each disorder (Figure S4F). We carried out competitive gene-set analysis via MAGMA due to its effectiveness in controlling for type-1 error in respect to confounding effects of gene size and gene density regarding linkage disequilibrium among SNPs in gene territory^41, 65^. The results from gene set analysis aligned with our previous observation, indicating that the combined effect size for having each of the five studied DGBIs was higher in enteric neurons and enteric glia (Figure 3D). Across both humans and adult datasets, this was primarily driven by functional chest pain and functional fecal incontinence for enteric neurons and functional diarrhea and functional fecal incontinence for enteric glia (Figure S4F).

As another unbiased method to test if enteric neurons are indeed the culpable GI cell type associated with DGBIs, we re-clustered the GI cell types based on their DGBI-associated transcript module scores (category 2). Using the cells from both the fetal and adult GI datasets, we created a Seurat object of cell by DGBI transcriptional scores for each of the five DGBI categories. Following the standard Seurat pipeline, the cells were clustered by their DGBI transcriptional scores revealing four populations (Figure 3E). Cluster 3 is associated with all DGBIs except for functional fecal incontinence, cluster 2 is associated with functional dyspepsia and functional fecal incontinence, cluster 1 is mildly associated functional diarrhea, and cluster 4 is associated with no DGBIs (Figure 3E-F). After resolving the representation of each GI cell type in each cluster, enteric neurons were most highly represented in cluster 3, replicating our previous findings that enteric neurons are highly associated with functional chest pain, functional diarrhea, functional dyspepsia, and functional dysphagia (Figure 3G). Notably, ICCs and myocytes make up most of cluster 2, which is associated with functional dyspepsia and functional fecal incontinence, again suggesting that multiple cell types may be affected by the molecular signatures of these DGBIs and contribute to the pathophysiology (Figure 3G). Finally, epithelial cells and enteric glia contribute mostly to clusters 1 and 4, respectively, suggesting that they have the lowest expression of DGBI-associated transcripts, since only cluster 1 was modestly associated with functional diarrhea (Figure 3G).

In summary, our analyses suggest that enteric neurons are highly associated with multiple DGBIs and therefore likely contribute to the pathogenesis and pathophysiology of the implicated disorders.

### DGBIs are associated with distinct enteric neuron neurochemical identities

Our expression-based analysis has identified that enteric neurons highly express molecular features associated with multiple DGBIs. However, enteric neurons consist of a multitude of unique neurochemical and functional subtypes, containing a neurochemical diversity comparable to the brain^13, 66^. Thus we hypothesized that enteric neurons do not all contribute to DGBIs in a nonspecific manner, but likely unique neurochemical subtypes drive each DGBIs pathology. In fact, the selective dysfunction of nitrergic neurons is previously shown to underlie GI disorders, such as achalasia, hypertrophic pyloric stenosis, and gastroparesis^67^. However, it remains unclear which enteric neuron subtypes may contribute to functional chest pain, functional diarrhea, functional dyspepsia, functional dysphagia, and functional fecal incontinence. To test this hypothesis and predict the neuronal populations associated with these DGBIs, we evaluated their association with the molecular features expressed in 21 neurochemical identities curated from four published human ENS transcriptomic datasets, including the two primary human datasets^14, 21^ and two human pluripotent stem cell-derived enteric neuron methods published by our group^22, 23^ (Figure 4A, Figure S5A). For neurotransmitters, we utilized our established two-step neurochemical identity-defining approach to identify all the cells that belong to six neurotransmitter identities: serotonergic (SRTNR), GABAergic (GABA), catecholaminergic (CATAMN), glutamatergic (GLUMT), cholinergic (CHOLN), and nitrergic (NITRG) neurons^22, 23^ (Figure 4B). For neuropeptides, we evaluated 15 identities that our group has defined previously based on the expression of calcitonin 2 (CTN-2), CART prepropeptide (CARTPP), cerebellin 2 (CERB-2), cholecystokinin (CCK), chromogranin A (CHRG-A), endothelin 3 (ET-3), galanin (GALAN), neuromedin U (NMDIN), neuropeptide Y (NP-Y), neuroexophilin 2 (NRXPH-2), proenkephalin (PRNKPH), secretogranin 3 (SGRN-3), somatostatin (SSTTN), substance P (SUB-P), and urocortin (UROCR)^23^ (Figure 4B).

**Figure 4:**
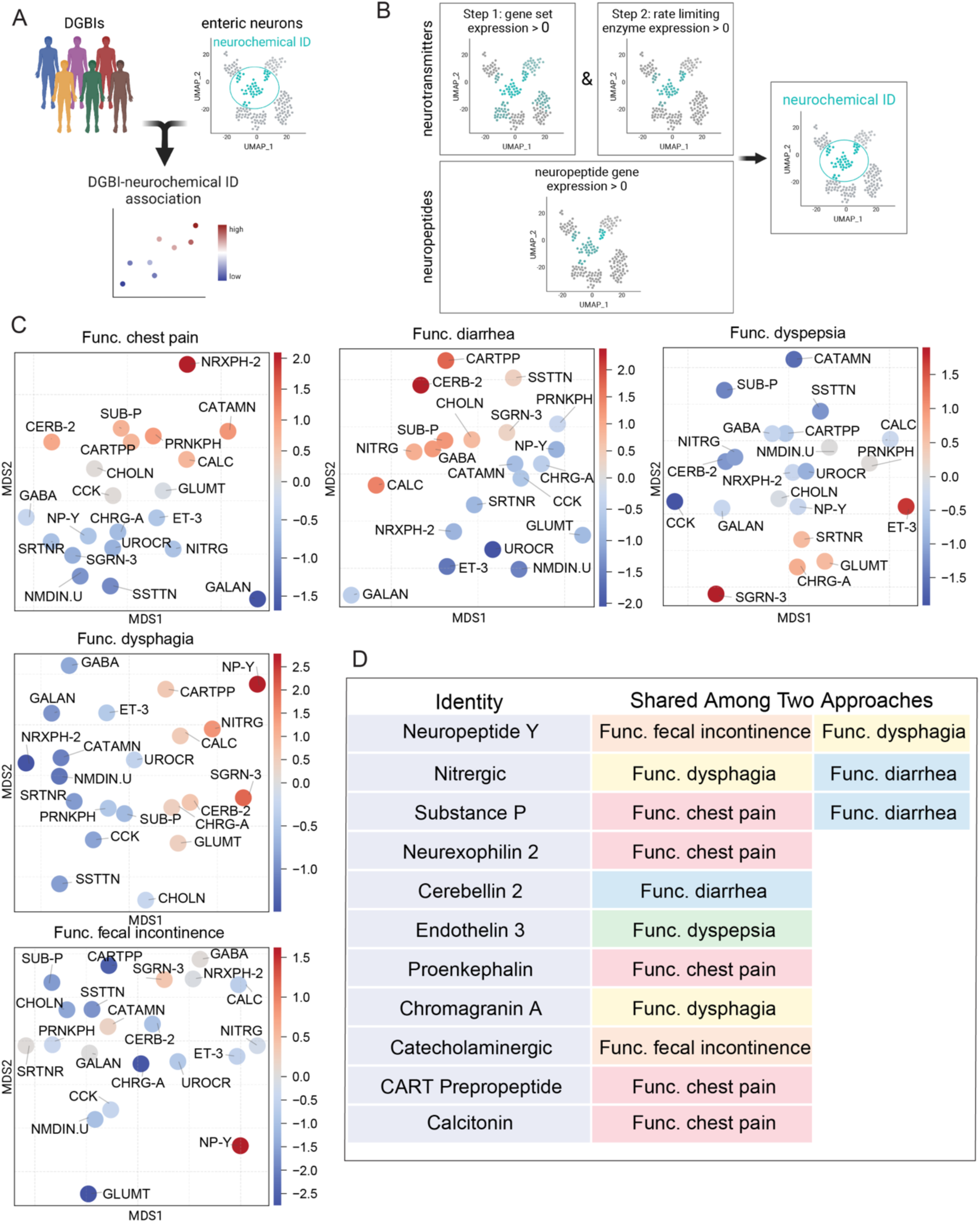
Expression and gene set association testing-based methods identify enteric neuron subtypes highly associated with each DGBI. **A)** Schematic illustration of the analysis method to identify enteric neuron subtypes associated with DGBIs. **B)** Schematic illustration of two-and one-step approaches to identifying neurotransmitter and neuropeptide subtypes in single cell transcriptomics datasets. **C)** Scatter plot showing the relationship between individual enteric neuron subtypes and their combined DGBI score based on module score analysis and DE association gene-set enrichment analysis. Distances are calculated via multidimensional scaling and disease scores are the average score of two methods. (Galanin: GALAN, Endothelin.3: ET-3, Nitrergic: NITRG, Dopamanergic: DOPAM, Glutamatergic: GLUMT, Urocortin: UROCR, Somatostatin: SSTTN, Chromogranin.A: CHRG-A, Calcitonin.2: CTN-2, Neurexophilin.2: NRXPH-2, Proenkephalin: PRNKPH, Neuromedin: NMDIN, CART.Prepropeptide: CARTPP, Substance.P: SUB-P, Neuropeptide.Y: NP-Y, Cholecystokinin: CCK, Cholinergic: CHOLN, Secretogranin.3: SGRN-3, Serotonergic: SRTNR, GABAergic: GABA, Cerebellin.2: CERB-2, Catecholaminergic: CATAMN, Calcitonin: CALC) **D)** List of the neuronal subtypes with high disease association scores in both analysis approaches and their corresponding diseases.

To investigate the association of the 21 neurotransmitter/neuropeptide identities with each of the five DGBIs, we tested the association of each identity’s differentially expressed genes leveraging each DGBIs transcript based association data from our GWAS. The results of these analyses for each of the four datasets were then averaged to generate an overall association score for each neurochemical identity with each DGBI (Figure 4B, Figure S5A,B). Each DGBI category had a distinct set of positively associated neuron subtypes. Cerebellin 2 and GABAergic neurons were mostly associated with functional chest pain, galanin, cerebellin 2; glutamatergic neurons were mostly linked with functional diarrhea; endothelin 3, glutamatergic, and calcitonin 2 were mostly associated with functional dyspepsia; secretogranin 3, cerebellin 2, and nitrergic neurons were mostly linked with functional dysphagia; and endothelin 3, glutamatergic, and calcitonin 2 neurons were mostly associated with functional fecal incontinence (Figure S5B).

As an orthogonal method to evaluate the link between enteric neuron subtypes and DGBIs, we assessed the expression of our curated DGBI gene (category 1 and 3) and transcript (category 2) lists (Table S5) in each of the 21 neurochemical identities across all four transcriptomic datasets. For each of the five DGBIs, we combined their respective genes and transcripts across the three categories and assessed their expression in each of the four transcriptomic datasets by module scoring. Averaging the DGBI scores for each neurochemical identity across all four datasets again revealed distinct neuron subtypes most positively associated with each DGBI, including neurexophilin 2 and catecholaminergic neurons for functional chest pain, CART prepropeptide, cerebellin 2, and somatostatin neurons for functional diarrhea, secretogranin 3 neurons for functional dyspepsia, and neuropeptide Y neurons for functional dysphagia and functional fecal incontinence (Figure S6). Furthermore, our curated list of published genes (category 3) revealed other unique enteric neuron subtype combinations associated with achalasia, constipation, and IBS (Figure S6)

Comparing the results of our gene set association testing method with our module score-based expression method, there were a number of observations for functional chest pain, functional diarrhea, functional dyspepsia, functional dysphagia, and functional fecal incontinence that were similar across both approaches (Figure 4C, Figure S5B, Figure S6). Both methods identified substance P, neurexophilin 2, proenkephalin, CART prepropeptide, and calcitonin neurons to be positively associated with functional chest pain; nitrergic, substance P, and cerebellin 2 neurons to be positively associated with functional diarrhea; endothelin 3 neurons to be positively associated with functional dyspepsia; neuropeptide Y, nitrergic, and chromogranin A neurons to be positively associated with functional dysphagia; and neuropeptide Y and catecholaminergic neurons to be positively associated with functional fecal incontinence (Figure 4C-D, Figure S5B, Figure S6). Notably, inhibitory motor neuron subtypes including, nitrergic and neuropeptide Y neurons were positively associated with multiple DGBI categories, further supporting the role of inhibitory motor neurons in GI disorders and diseases, as observed for achalasia, gastroparesis, hypertrophic pyloric stenosis, Hirschsprung’s disease, and Chagas’ disease^67, 68^.

By our two pronged gene set association testing and module score-based expression approaches, we identified distinct enteric neuron subtypes that consistently show high associated with each of the five DGBIs. Overall, this suggests that distinct enteric neuron subtype combinations are mechanistically linked with DGBIs and may underlie distinct DGBI pathophysiologies.

### Network analysis reveals DGBI relevant molecular pathways

We next aimed to identify the genes driving the association between the neuronal subtypes and each of the five disorders. We leveraged the DGBI associated genes, curated from our SNP and gene-based genome wide association testing as well as the genes identified from previously published GWAS studies (Table S6) and filtered the genes based on their expression in enteric neuron subtypes across all four transcriptomic datasets. To explore if the commonly expressed genes associated with each DGBI are involved in specific molecular pathways, we utilized the STRING physical interaction database to perform a protein-protein interaction network analysis. Each DGBI formed its own unique set of networks, with functional diarrhea containing one core network of more than two nodes, functional chest pain, functional dyspepsia, and functional dysphagia containing two core networks of more than two nodes, and functional fecal incontinence containing three core networks of more than two nodes (Figure 5A). The networks are largely shared across the implicated enteric neuron subtypes, with very few genes being specific to one or more subtypes (Figure 5A). For example, in the functional chest pain network, *TMEM37* and *PTPN4* are only expressed in CART prepropeptide, neurexophilin 2, and substance P neurons, while calcitonin and proenkephalin enteric neurons lack these network nodes (Figure 5A). However, these proteins satellite the core networks, which are conserved across the implicated cell types, suggesting that the same molecular mechanisms may drive DGBI pathology in the enteric neurons subtypes associated with the DGBIs (Figure 5A).

**Figure 5:**
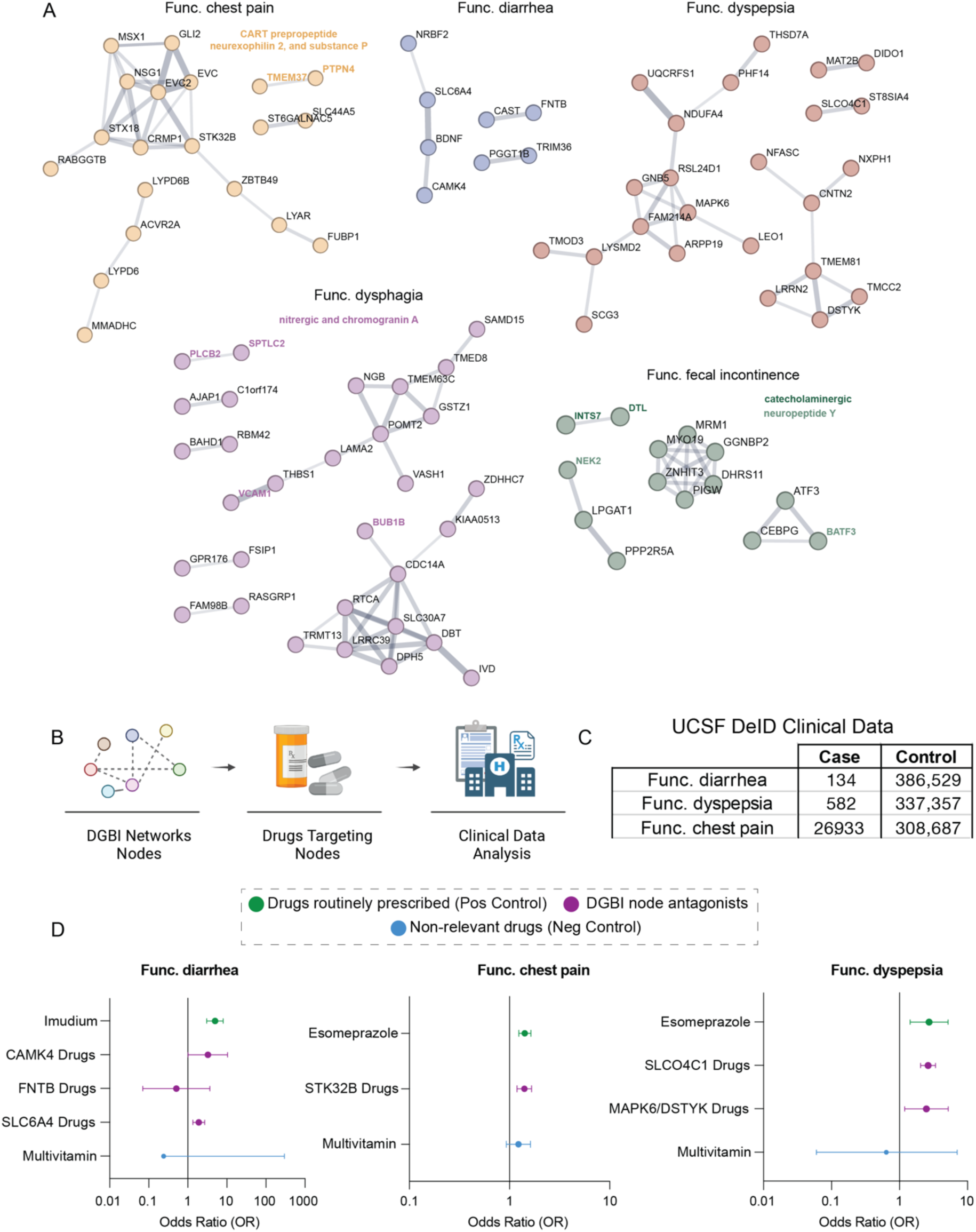
Protein-protein interaction analysis identifies proteins and protein pathways linked with DGBI risk. **A)** STRING protein-protein interaction networks formed from the DGBI associated genes. Protein nodes specific to an enteric neuron subtype are highlighted. The minimum required interaction score was set to 0.4, reflecting medium confidence interactions. **B)** Schematic illustration of the analysis pipeline to examine links between medications that target the network protein nodes and DGBIs in clinical records. **C)** Number of cases and controls UCSF extracted from the de-identified clinical database. **D)** Multivariate logistic regression analysis of the odds of DGBIs in patients treated with network-targeting drugs as well as commonly prescribed drugs for each DGBI as positive controls and multivitamin as negative control.

The core of functional chest pain’s largest network contains three proteins involved in the hedgehog signaling pathway: evc ciliary complex subunit 2 (EVC2), evc ciliary complex subunit 1 (EVC), and GLI family zinc finger 2 (GLI2)^69^, suggesting a novel role of hedgehog signaling in potentiating functional chest pain in enteric neurons (Figure 5A). These nodes, in addition to serine/threonine kinase 32B (STK32B) and MSX homeobox 1 (MSX1), are associated with developmental defects, including Ellis-van Creveld syndrome^70, 71^, where short stature and heart malformations are a common disease manifestation and a patient’s cause of death is often due to heart failure^72^. Notably, functional chest pain often precedes a patient’s diagnosis of ischemic heart disease^73^, further supporting the role of these proteins and developmental pathways in potentiating functional chest pain symptoms. Importantly, in our GWAS study design, patients with a history of cardiac diseases with manifestations of chest pain were excluded. Thus, in our study, the identification of these genetic variants relating to hedgehog signaling are not due to chest pain with cardiac etiology. Instead, these results point to a novel role of hedgehog signaling in potentiating functional chest pain in enteric neurons.

Intriguingly, brain derived neurotrophic factor (BDNF) and serotonin transporter (SLC6A4) are at the centeri of functional diarrhea’s network. These factors are implicated in psychiatric disorders, including post-traumatic stress disorder, bipolar disorder, and depression^74^ (Figure 5A). Previous reports have found a strong association between GI diseases and anxiety and stress-related psychiatric disorders^24, 75^. In fact, recent work has found that these disorders may arise independently due to conserved mechanisms found both in the brain and the gut rather than one causing the other^24^. Our analysis supports this observation, as we found that psychiatric disorder-associated genes are highly expressed in the enteric neuron subtypes associated with functional diarrhea, specifically nitrergic, cerebellin 2, and substance P neurons (Figure S6). Mechanistically, the enrichment of neurotransmission-related ontology gene sets in our pathway analysis and the representation of neuronal function-related genes here, further suggests neurotransmission-related dysfunction in nitrergic, cerebellin 2, and substance P neurons may underlie functional diarrhea (Figure 3A and Figure 5A). Furthermore, targeting these dysfunctional neurons may offer a therapeutic benefit to functional diarrhea patients, a unique mechanism, given that available antidiarrheal drugs do not target neurons and are not compatible with chronic usage^76^.

Similar to functional diarrhea, the functional dyspepsia network also contains proteins essential for neurotransmission (Figure 5A). Neurofascin (NFASC), contactin 2 (CNTN2), and ST8 alpha-N-acetyl-neuraminide alpha-2,8-sialyltransferase 4 (ST8SIA4) are protein components in neuronal axons, with NFASC and CNTN2 functioning specifically the nodes of Ranvier^77, 78^. Furthermore, G protein subunit beta 5 (GNB5) and cAMP regulated phosphoprotein 19 (ARPP19) are known to be involved in dopamine signaling^79, 80^. Besides supporting the role of enteric neurons in functional dyspepsia, these neuron-specific proteins suggest defects in neuronal processes such as neurite extension, axonal guidance, synaptogenesis, neuronal plasticity, and neurotransmission may underlie functional dyspepsia pathophysiology. To date, the mechanisms underlying functional dyspepsia are poorly understood and it remains heavily debated if functional dyspepsia should be considered a subcategory of gastroparesis or vice versa since they share overlapping symptoms including delayed gastric emptying, which is a core clinical feature in gastroparesis^81^. Furthermore, like functional diarrhea, functional dyspepsia is largely managed through the use of medications such as proton pump inhibitors and histamine-2 receptor antagonists. These treatments help to manage symptoms but based on our study do not target the underlying mechanism or the implicated enteric neuron subtype (endothelin 2 neurons)^81^. Thus, by uncovering the high confidence enteric neuron subtype and molecular features specific to functional dyspepsia, our study offers new opportunities for a better understanding of the molecular mechanisms of this intractable and poorly understood DGBI.

In order to determine the clinical relevance of these protein interaction networks, we identified all FDA approved drugs that antagonize or inhibit the nodes of each disease network (full list of drugs can be found in Table S7). Next, we leveraged patient medical record data from the UCSF De-Identified Clinical Data Warehouse to evaluate the association of the node targeting therapies with DGBI occurrence. We reviewed the database for each disease separately, and filtered the diseases and conditions with similar inclusion and exclusion criteria as we used previously in the UKBB (Table S1). In the filtered datasets, we defined cases and controls for each DGBI and assessed whether the prescription of node targeting drugs is associated with a higher or lower incidence of DGBI. Functional dysphagia and functional fecal incontinence protein network nodes did not have sufficient approved drugs or prescription history. For the other three DGBIs, a commonly prescribed drug for the indication was used as a positive control and a multivitamin drug served as a negative control drug to validate the methodology. By using a multivariate logistic regression analysis adjusted for age, sex, smoking and BMI, we examined the associations of the drug families and the positive and negative controls with each corresponding GI disorder. There was a significant association between drugs that either antagonize or inhibit SLC6A4 for functional diarrhea, SLCO4C1, MAPK6 and DSTYK for functional dyspepsia, and STK32B for functional chest pain (Figure 5D). These results provide support for the role of these proteins in DGBIs, suggesting that in addition to our previous observations that genetic variants in their corresponding genes may predispose patients to developing DGBIs, their function at the protein level can also be a contributing factor. Furthermore, our study identified that drugs targeting these networks are significantly associated with DGBI pathology. Nominating these networks to be studied in DGBIs in prospective well controlled studies in the future to find effective treatments.

Altogether, our study presents a framework for uncovering the cells, cell types, and proteins involved in DGBIs by leveraging large-scale human genetic, transcriptomic, and medical record data (Figure 6). Here we validated the link between functional chest pain, functional diarrhea, functional dyspepsia and their corresponding DGBI networks, observing a significantly increased risk of DGBI symptoms when patients were prescribed drugs that antagonize proteins in the DGBI network. These results not only uncovered high confidence proteins and protein pathways underlying DGBIs but also offer a strategy for future drug discovery efforts for these intractable conditions that have seen little to no therapeutic advancement in recent years.

**Figure 6:**
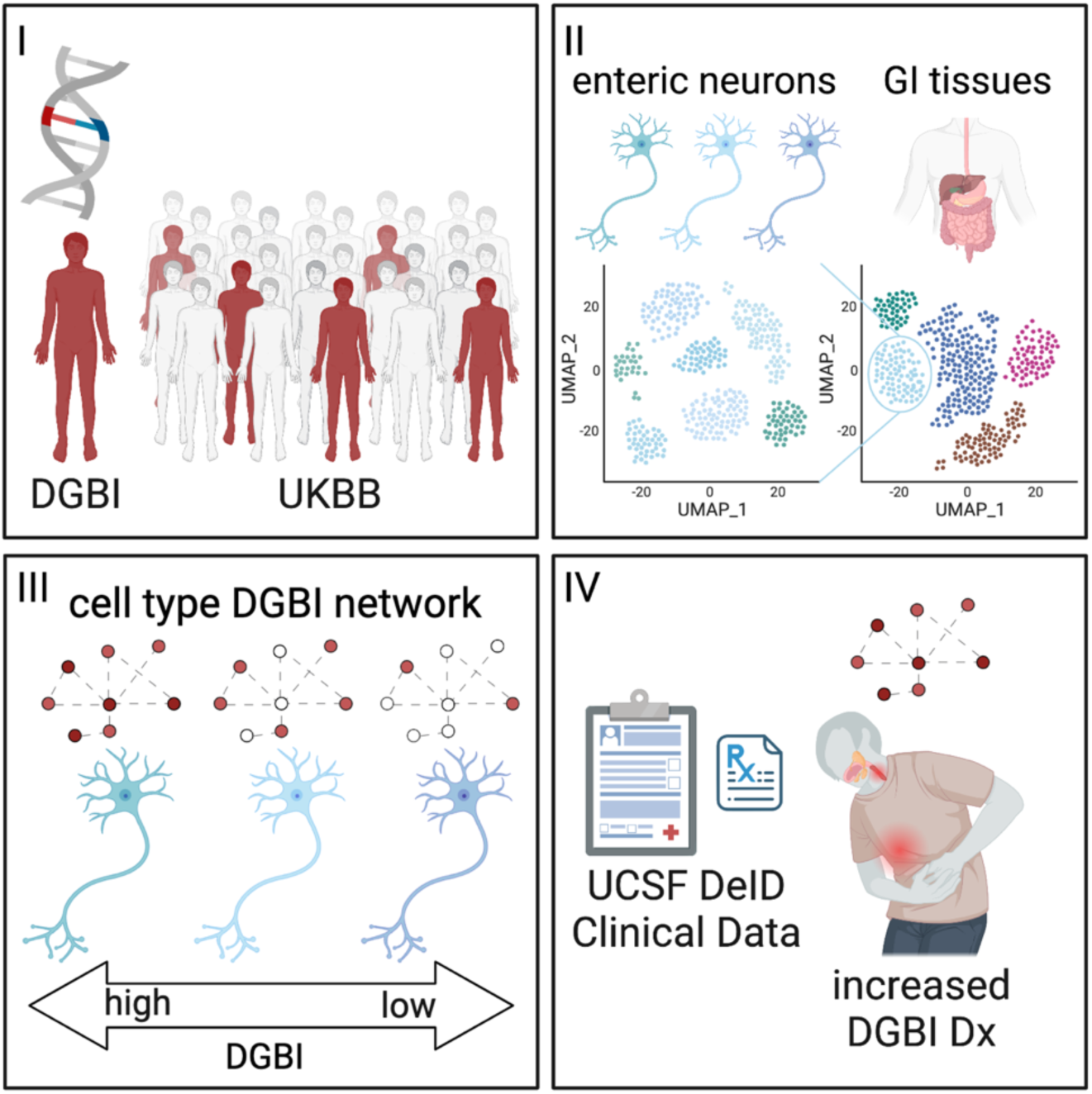
Schematic illustration of the study framework.

## Discussion

DGBIs are extremely common and historically difficult to manage. This is largely because the cellular and molecular mechanisms underlying DGBIs remain poorly understood and understudied. Here we leveraged the UKBB database and applied strict inclusion and exclusion criteria to perform GWAs on five DGBIs: functional chest pain, functional diarrhea, functional dyspepsia, functional dysphagia, and functional fecal incontinence. To ensure easy accessibility for the research community, we have created an online tool at BBNbrowser.com, which allows researchers to find DGBI disease association for any gene of interest.

Our comprehensive and stringent study design identified significant association between the rs975577 variant (in the *MRM1* gene) and the rs2081046 variant for functional fecal incontinence. Functional and structural annotations of potential variants with a less strict threshold (1e-6) revealed in total 162 annotated genes for the five DGBIs studied. *S1PR1* is mapped to functional dysphagia-associated variants and has been previously associated with IBS. Similarly, functional chest pain SNPs were annotated to *GLI2* and *CRMP1* and were previously identified to be linked with functional dysphagia and Hirschsprung’s disease. Therefore, these genes might be suitable candidates to provide insight into the pathophysiology behind multiple DGBIs. Through the translation of SNP association data into gene masks, we conducted genome-wide gene-based association testing and identified two significant genes for functional dysphagia, *RBM42* (p-adj 0.02) and *LAMA2* (p-adj 0.04) highlighting their potential as promising candidates for further investigation.

To obtain further insight into potential genetic mechanisms, we filtered genes based on their z-score of association and curated a list of 159 genes. Interestingly, GPR176 for functional dysphagia, as well as RABGGTB and ZNF618 for functional chest pain, are present in our annotated genes for SNP hits associated with the same diseases, thus supporting the potential association of these genes. Notably, *BACE2*, which is associated with the combined DGBI phenotype in our analysis, has previously been linked to Hirschsprung’s disease^42, 82^. These genetic associations were highly enriched in pathways relating to nervous system functions including neurotransmission-related gene sets, heavily implicating the ENS and enteric neurons in the pathogenesis of these DGBIs. Notably, loss or dysfunction of enteric neurons has been observed in other GI disorders, such as achalasia, hypertrophic pyloric stenosis, and gastroparesis^67^. Thus, the enrichment of our unbiasedly identified genetic features of DGBIs with neuron-specific pathways further supports the mechanistic connection of the ENS with DGBI pathophysiologies.

Our study further highlights the role of enteric neurons in DGBIs. Leveraging multiple human single cell transcriptomic datasets to evaluate the association of GI cell types with our five DGBI categories, we found enteric neurons to be highly associated with the DGBIs. Focusing in on the enteric neurons, we applied two expression-based association methods to identify the enteric neuron subtypes significantly correlated with each DGBI. This analysis revealed novel cell type-DGBI associations including substance P, neurexophilin 2, proenkephalin, CART prepropeptide, and calcitonin neurons for functional chest pain, nitrergic, substance P, and cerebellin 2 neurons for functional diarrhea, endothelin 3 neurons for functional dyspepsia, neuropeptide Y, nitrergic, and chromogranin A neurons for functional dysphagia, and neuropeptide Y and catecholaminergic neurons for functional fecal incontinence. Furthermore, these implicated cell types similarly express the DGBI-associated genes whose proteins form protein-protein interaction networks. These high confidence DGBI mechanism predictions suggest that each DGBI may be driven by dysregulation of molecular pathways shared across the implicated enteric neuron subtypes. Future studies leveraging *in vitro*, *ex vivo*, and *in vivo* model systems are warranted to evaluate and validate the functional role of the cell types and proteins we have found to be associated with each DGBI.

Finally, our study reveals several important implications for the understanding and management of DGBIs as the identification of specific genetic risk factors, proteins, and implicated cell populations may help to improve the diagnosis, prognosis, and management of these conditions. For example, in our medical record analysis we evaluated the prevalence of DGBIs in patients prescribed medications that inhibit or antagonize proteins in our DGBI protein-protein interaction networks. This analysis revealed multiple approved drugs associated with increased risk of DGBIs, including SLC6A4 targeting drugs for functional diarrhea, SLCO4C1, MAPK6, and DSTYK targeting drugs for functional dyspepsia, and STK32B targeting drugs for functional chest pain. It is unsurprising that cancer drugs, including MAPK6, DSTYK, and STK32B inhibitors cause DGBI symptoms, since GI toxicity, including nausea, vomiting, diarrhea, and constipation, is the most common adverse reaction to cancer therapy^83^. However, the prevalence of DGBIs symptoms that correspond with the DGBIs we studied is notable. For example, neratinib (a MAPK6 inhibitor) and sunitinib (a DSTYK inhibitor) are known to cause dyspepsia in some patients, while antidepressants like citalopram (a SLC6A4 antagonist) can cause diarrhea, as reported in their packaging inserts. Furthermore, for non-antineoplastic agents such as the SLCO4C1 targeting drugs digoxin, liothyronine, and methotrexate, dyspepsia symptoms including abdominal distress, cramps, and pain are listed as adverse reactions in their packaging inserts, highlighting the potential of our large data analysis to identify the underlying DGBI not detected or reported during clinical trials. These results also could be explained by the progressive and non-specific nature of DGBIs. During a short-term clinical trial, it may be infeasible to diagnose a DGBI appropriately, but our analysis is able to identify the strong link between these drugs, their DGBI-inducing symptoms, and their eventual DGBI diagnosis. Ultimately, these results identify potential drugs to be contraindicated for the corresponding DGBI, due to the risk of exacerbating the condition. Overall, further research is needed to fully understand the interplay between the genetics, proteins, and implicated cell populations in the development and progression of DGBIs in order to determine the most effective ways to manage these conditions.

Our study offers a new framework for leveraging a large-scale database to assess genetic associations with five DGBIs. The UKBB is an invaluable resource as one of the largest and most comprehensive biobanks. However, as databases, such as the UKBB, containing genetic and health record data grow, efforts must be made to enhance the inclusion of individuals from diverse ancestral backgrounds. Currently the UKBB is primarily composed of Caucasian British participants and is not fully representative of the global population. This must be recognized as a source of bias when interpreting findings from UKBB studies, since genetic variation and disease susceptibility vary across different populations worldwide. Incorporating a diverse range of populations into similar studies and cross-validating the results will be crucial to enhance the generalizability of the findings beyond the specific demographics represented in the UKBB.

Furthermore, our use of extensive exclusion criteria has reduced the sample size for each DGBI category, which could introduce potential biases. Future independent studies with similar criteria in a different database, are needed to further validate these findings. Our medical records analysis examining drugs targeting protein networks is retrospective and cross-sectional in nature, and hence conclusions should be verified with prospective and carefully controlled studies.

In conclusion, our results provide evidence for a significant genetic correlation between DGBIs and specific enteric neuron subtypes, implicating their involvement in DGBI pathophysiology. Overall, this study highlights the promise of using human genetic and medical record data in tandem with human transcriptomic data to investigate and reveal novel mechanisms of complex diseases.

## Data Availability

All data produced in the present work are included in the manuscript and available at bbnbrowser.com.

http://bbnbrowser.com/

## Acknowledgements

This research has been conducted using the UK Biobank Resource under Application Number 61335. The authors acknowledge the use of the UCSF Information Commons computational research platform, developed and supported by UCSF Bakar Computational Health Sciences Institute. This publication was supported by the National Center for Advancing Translational Sciences, National Institutes of Health, through UCSF-CTSI Grant Number UL1 TR001872. Its contents are solely the responsibility of the authors and do not necessarily represent the official views of the NIH. The work was generously supported by the NIH Director’s New Innovator Award (DP2NS116769) and the National Institute of Diabetes and Digestive and Kidney Diseases (R01DK121169) awarded to F.F.

## Authors Contributions

A.M. Defined case and control cohort and disease definitions in UK Biobank, patient characterization and statistical analyses, co-morbidity analysis. Performed genome wide association studies, post-GWAS studies and Gene-based association studies. Performed gene set analysis of DE genes for GI cell types and neurochemical identities. Designed and completed UCSF de-identified clinical data analysis. Built the BBNBrowser website. Wrote the manuscript.

M.N.R. Performed pre-ranked gene set enrichment analysis. Designed and completed transcriptomic analyses including calculating cell type transcriptional signatures, DGBI transcriptional scoring of GI cell types and enteric neuron subtypes, re-clustering analysis of GI cell types by DGBI module scores, and the protein-protein interaction network analysis. Wrote the manuscript.

R.M.S. Assisted with single cell transcriptomic analyses including curating the published datasets.

A.C. Assisted with single cell transcriptomic analyses including curating the published datasets.

Z.G. Assisted with the preparation of UK Biobank data, data access, disease definitions, and GWAS.

J.W. Assisted with UK Biobank data preparation and genomic data cleaning.

F.F. Designed and conceived the study, supervised all analyses, and wrote the manuscript.

---

**Supplementary Figure 1:**
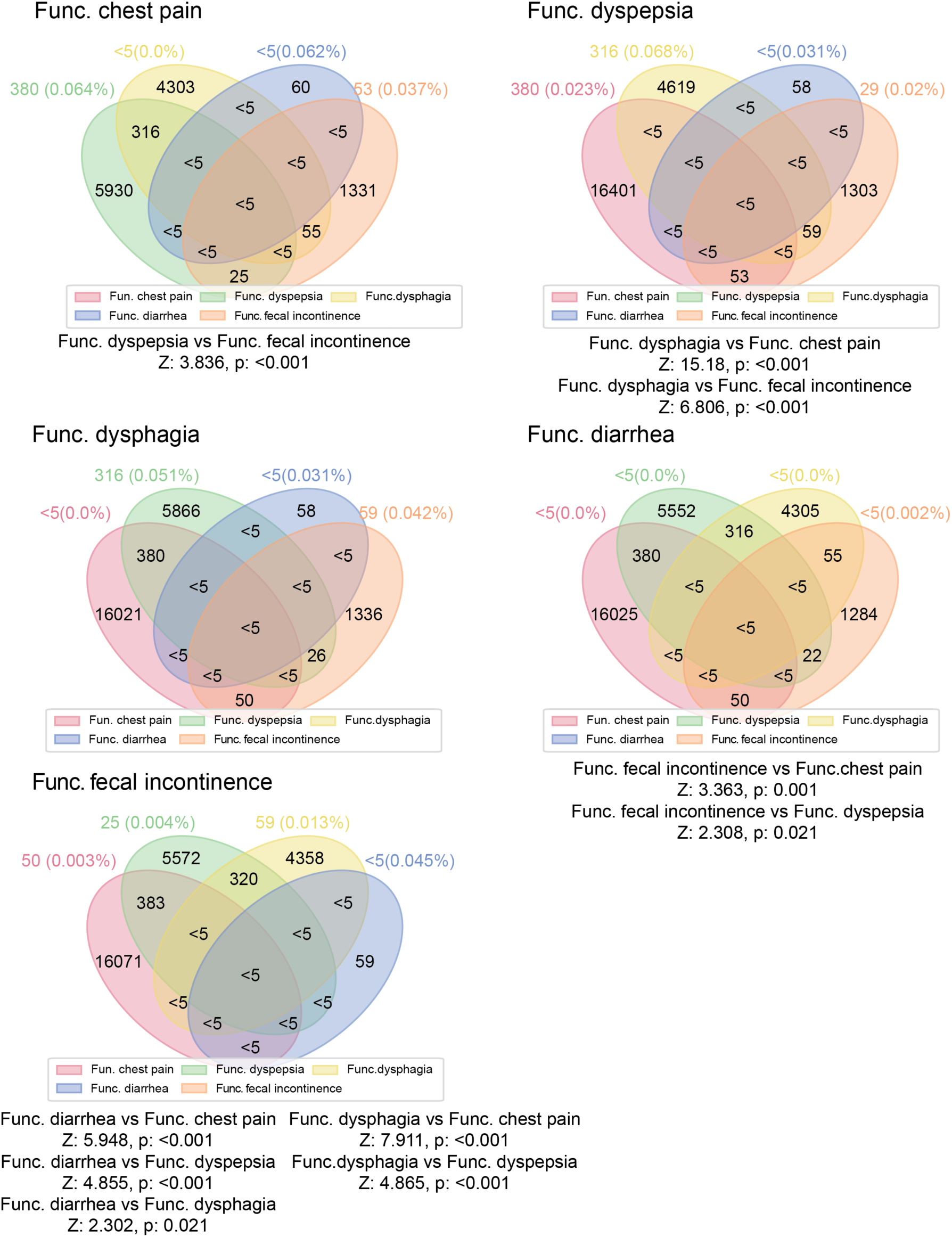
Co-morbidity analysis of DGBIs. Venn diagram illustratation the co-morbidity analysis, in which one disease serves as an index disease (indicated at the upper left of each diagram) and the number of individuals shared between the index disease and the other diseases is compared. We used the two-proportion Z test to determine which diseases are significantly more comorbid with the index disease. Results with p-values <0.05 in each round are shown beneath each diagram.

**Supplementary Figure 2:**
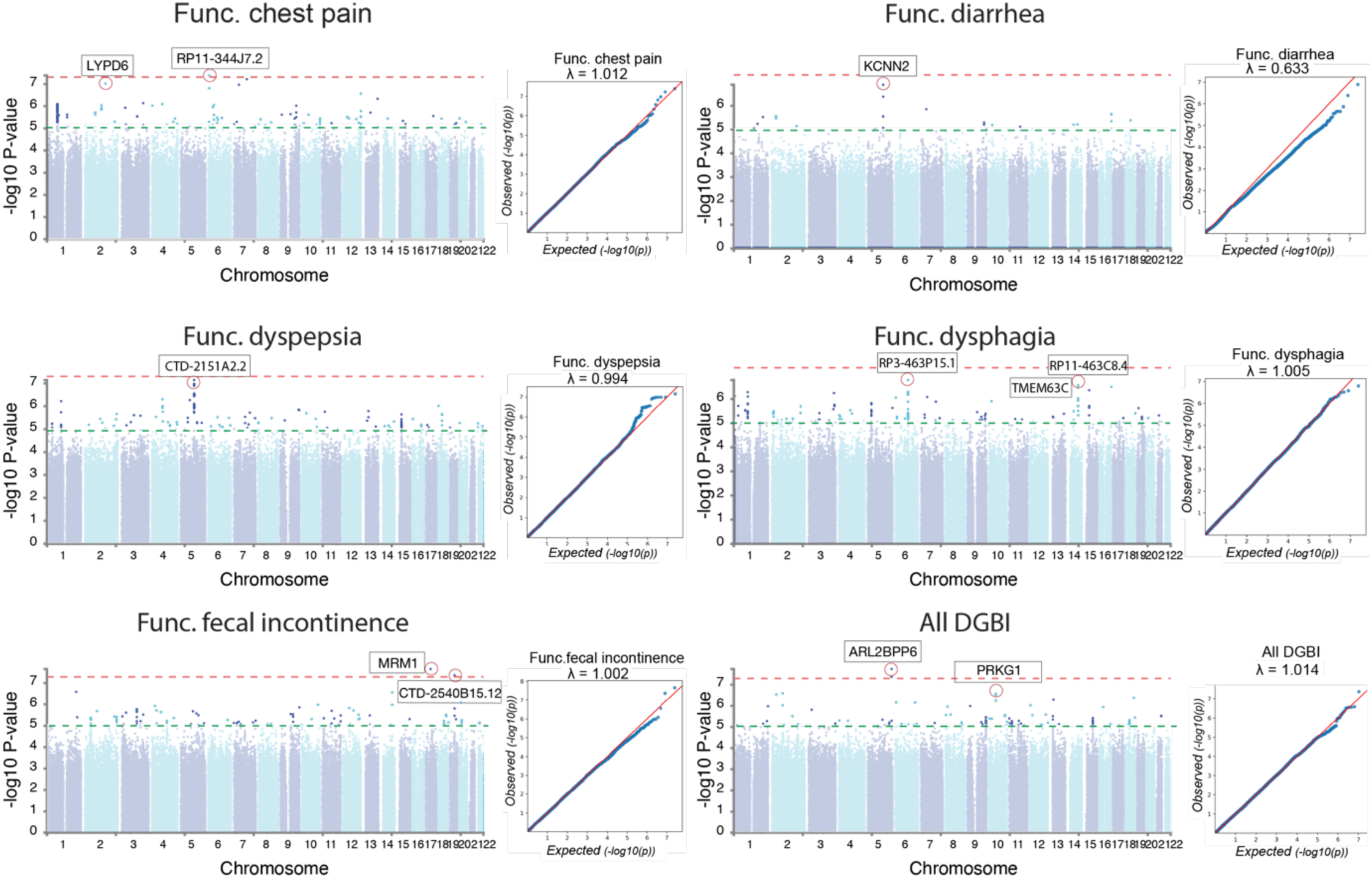
Identification of significant DGBI associated SNPs. Manhattan plots and QQ plots from 5 DGBIs and one combined GWAS. The closest genes are highlighte for the most promising SNPs.

**Supplementary Figure 3:**
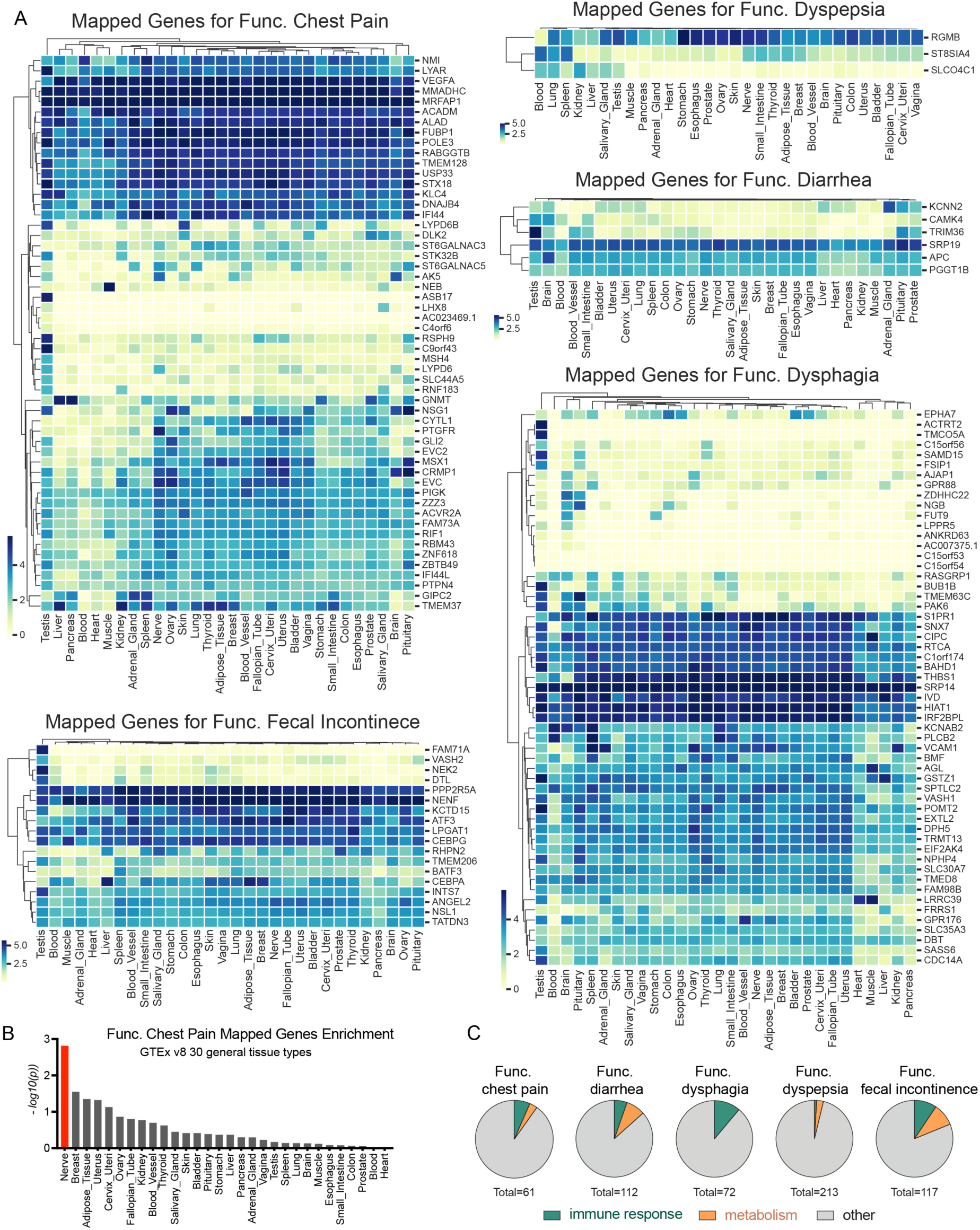
Expression patterns of DGBI associated genes. **A)** Heatmaps representing the expression levels of DGBI associated genes across 30 general tissue types based on GTEx v8 30. **B)** Comparison of enrichment of genes associated with functional chest pain across differentially expressed genes in various tissue types. Significant enrichment at Bonferroni corrected P-value ≤ 0.05 are colored in red. **C)** Pie charts showing the proportion of immune response and metabolism gene sets significantly enriched among DGBI associated genes.

**Supplementary Figure 4:**
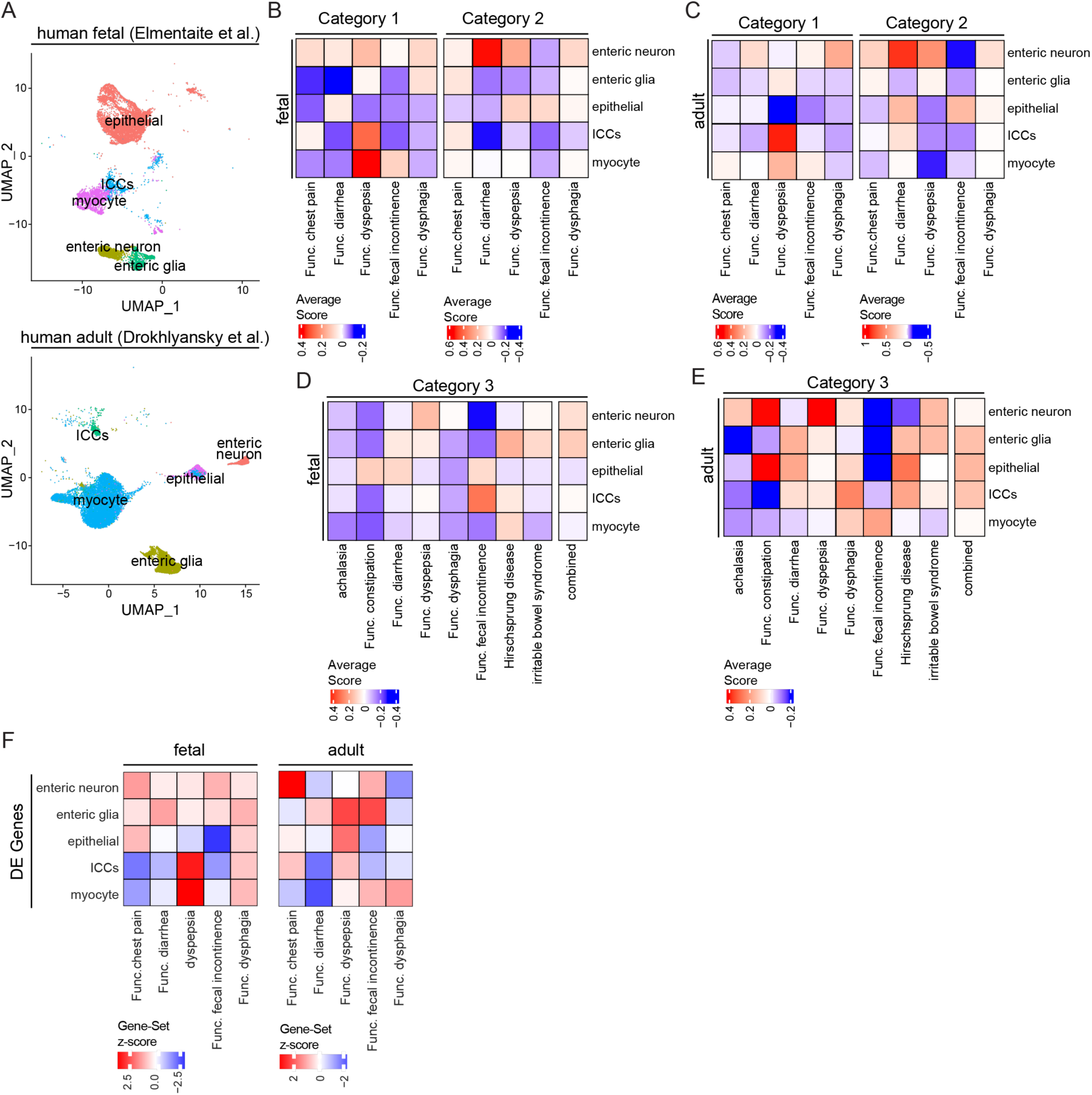
Identification of the GI cell types associated with DGBIs. **A)** UMAPs of the enteric neurons, enteric glia, epithelial cells, ICCs, and myocytes in the human fetal intestine scRNA-seq dataset (top) and the human adult colon snRNA-seq dataset (bottom). **B, C**) Heatmaps of the average module scores of DGBI associated molecular features in the human fetal and adult GI cell types for each DGBI category using significantly associated transcripts from gene-based testing (category 1) and significantly associated genes from SNP-based testing (category 2). **D, E**) Heatmaps of the average module scores of DGBI associated molecular features in the human fetal and adult GI cell types for each DGBI category using significantly associated genes from published DGBI GWAS studies (Category 3). **F**) Heatmap of the DGBI association scores for clusters of human fetal and adult GI cells based on gene-set enrichment analysis.

**Supplementary Figure 5:**
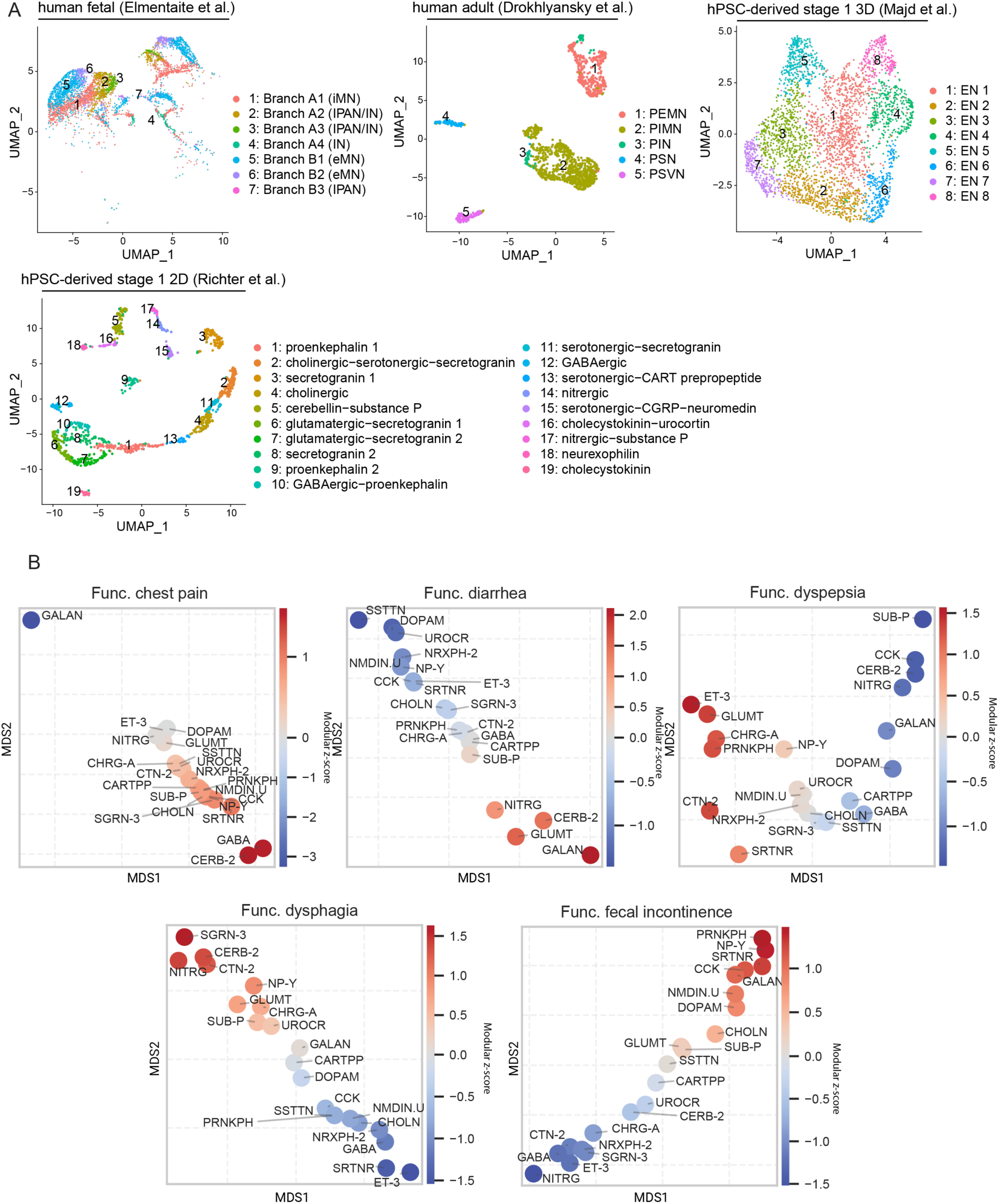
Identification of enteric neuron subtypes associated with DGBIs. **A)** UMAPs of the annotated enteric neurons subtypes in the human fetal intestine scRNA-seq dataset, the human adult colon snRNA-seq dataset, the hPSC-derived stage 1 ganglioid snRNA-seq dataset, and the hPSC-derived stage 1 2D scRNA-seq dataset. **B)** Scatter plot showing DGBI scores of enteric neuron subtypes based on gene-set enrichment analysis of subtype-specific DE genes. Distances are calculated via multidimensional scaling and values are the average of disease association z-scores across datasets. (Galanin: GALAN, Endothelin.3: ET-3, Nitrergic: NITRG, Dopamanergic: DOPAM, Glutamatergic: GLUMT, Urocortin: UROCR, Somatostatin: SSTTN, Chromogranin.A: CHRG-A, Calcitonin.2: CTN-2, Neurexophilin.2: NRXPH-2, Proenkephalin: PRNKPH, Neuromedin: NMDIN, CART.Prepropeptide: CARTPP, Substance.P: SUB-P, Neuropeptide.Y: NP-Y, Cholecystokinin: CCK, Cholinergic: CHOLN, Secretogranin.3: SGRN-3, Serotonergic: SRTNR, GABAergic: GABA, Cerebellin.2: CERB-2, Catecholaminergic: CATAMN, Calcitonin: CALC)

**Supplementary Figure 6:**
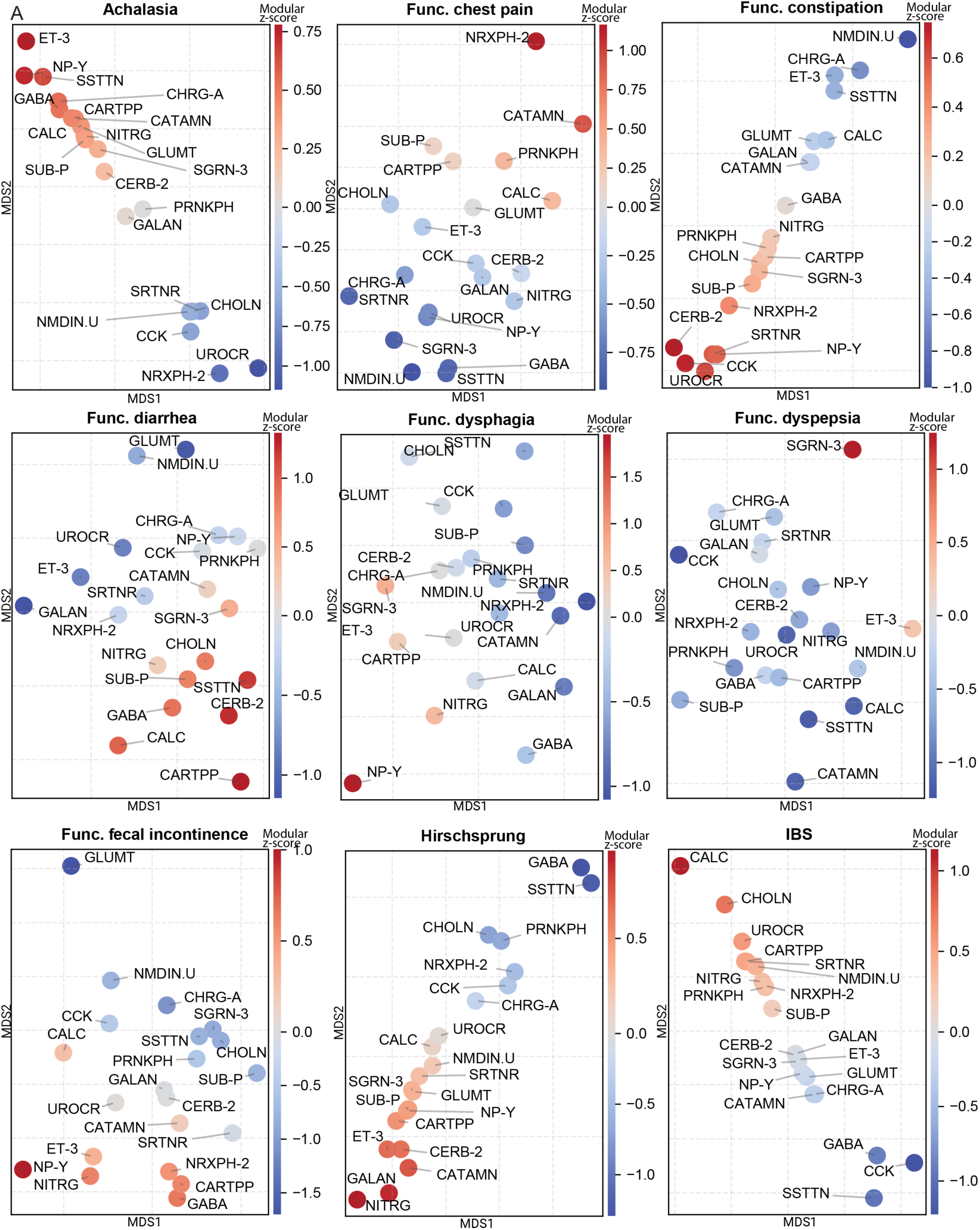
Identification of enteric neuron subtypes that express DGBI associated transcripts. Scatter plot showing DGBI scores of enteric neuron subtypes based on module scoring of DGBI signatures. Distances are calculated via multidimensional scaling and values are the average of DGBI modular scores across neuronal datasets. (Galanin: GALAN, Endothelin.3: ET-3, Nitrergic: NITRG, Dopamanergic: DOPAM, Glutamatergic: GLUMT, Urocortin: UROCR, Somatostatin: SSTTN, Chromogranin.A: CHRG-A, Calcitonin.2: CTN-2, Neurexophilin.2: NRXPH-2, Proenkephalin: PRNKPH, Neuromedin: NMDIN, CART.Prepropeptide: CARTPP, Substance.P: SUB-P, Neuropeptide.Y: NP-Y, Cholecystokinin: CCK, Cholinergic: CHOLN, Secretogranin.3: SGRN-3, Serotonergic: SRTNR, GABAergic: GABA, Cerebellin.2: CERB-2, Catecholaminergic: CATAMN, Calcitonin: CALC)

## Supplementary Tables

Table S1: Disease specific inclusion and exclusion codes.

Table S2: Cohort Characteristics.

Table S3: Top SNPs and functional/structural annotations.

Table S4: List of DGBI associated genes generated through gene-based association testing.

Table S5: Curated gene lists from previously published GWAS, SNP-mapped genes, and gene lists filtered by Z-score.

Table S6: List of genes associated with functional chest pain, functional diarrhea, functional dyspepsia, functional dysphagia, and functional fecal incontinence.

Table S7: Full list of drugs targeting DGBI network nodes used in clinical data analysis.

Table S8: List of genes used to define neurochemical identities.

## Methods

### Phenotype Definition

The UK Biobank (UKBB) (http://www.ukbiobank.ac.uk) is a large prospective database in which approximately 500,000 people gave their consent for comprehensive health, lifestyle, demographic, biometric and genetic data to be accessed in the UK Biobank.^14^ This study utilizes data from UKBB with registration number 61335. To identify cases for each DGBI, we used inpatient ICD-10 diagnoses and self-report conditions. The cases were identified using inpatient ICD diagnoses and self-report conditions (UKBB datafield 20002, 41201, 41202, 41204, 41270). By assessing a detailed past medical history of past surgeries and cancer histories, as well as a self-report and ICD diagnosis codes (previous data fields along with UKBB data fields 41200, 41210, 41272, 20004, 40006, and 2000), participants with any condition with potential similar manifestations were identified and excluded from cohorts. An extensive list of codes and inclusions and exclusions for each disease can be found in Table S1. For each disease, a control group is made up of the remaining individuals. DGBI cases were combined into a mixed phenotype called “All DGBI;” cases were defined as individuals with one or more diseases in their records, and controls were defined as individuals without any of these diseases or the combined list of exclusions in their records. Disease definitions and corresponding analysis and visualization were performed in python (version 3.8.8) using Pandas (version 1.4.1), NumPy (version 1.22.2), statsmodels (version 0.13.2) and seaborn (version 0.11.2) and in R (version 4.2.0) using gtsummary (version 1.6.1) and nVennR (version 0.2.3)^84^.

### Comorbidity analysis

We performed a stepwise comorbidity analysis by selecting one disease as the index group in every round. We determined the number of individuals in both the index disease group and each of the other disease groups (Figure S1). Subsequently, we performed two-proportion Z-test pairwise comparisons to determine which disease groups exhibited a higher comorbidity rate with the index disease (Figure S1). A similar approach has been employed in previous studies involving IBS, IBD, GORD, and PUD^43^.

For example, when disease D_i_ served as the index disease, a significant result between D_1_ vs D_2_ (with Z=3.5, p<0.001) indicates that D_i_ has higher comorbidity with D_1_ than D_2_.

We computed a “distance (d)” factor between all diseases using these z-score values. For example, we approximate the distance between D_i_ and D_2_ or d(D_i_-D_2_) by multiplying the distance between D_i_ and D_1_ or d(D_i_-D_1_) by 3.5.

> d(D_i_-D_!_) = 3.5*d(D_i_-D_”_)

We performed these calculations for all significantly comorbid disease pairs (p<0.05, Figure S1). To visualize all comorbidities relative to each other, we projected the results along a unidimensional comorbidity line where the diseases are ordered according to their relative distance.

In the example above, the linear projection D_1_-D_2_-D_i_ is excluded and there are two remaining possibilities: D_i_-D_1_-D_2_ or D_1_-D_i_-D_2_. If d(D_1_-D_!_) < d(D_i_-D_!_) the diseases will be ordered as D_i_-D_1_-D_2_ but if d(D_1_-D_!_) > d(D_i_-D_!_) they will be ordered as D_1_-D_i_-D_2_.

We then calculated the “relative distance (relative d)” of Di-D1 to the total distance:

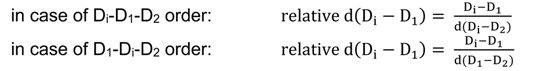

Finally, the relative distances were scaled to the most distant diseases (here Func. chest pain and Func. diarrhea) so the total relative distances sum up to 1 (Figure 1E).

### Genotype and Sample Quality Control

Genotype data were downloaded using UKBB’s ’gfetch’ tool and SNP quality controls were performed using PLINK^85, 86^ recommended procedures (SNPs with missingness of 0.05, Hardy Weinberg equilibrium test of 1e6 and Minor allele frequency threshold of 0.01). Participants whose self-reported sex did not match their genetically determined sex, kinship relatedness of >10 and only participants who identified as white British ancestry were included in the discovery dataset. Participants with self-reported sex that did not match their genetically determined sex, those with kinship relatedness of greater than 10 and those who identified as non-white British ancestry were excluded from the discovery dataset.

### Genome Wide Association Study (GWAS) Analysis

REGENIE v3.1.1 was used to conduct the GWAS analysis using a Firth bias-corrected logistic regression^37^. We adjusted the model for age, age squared, sex, age multiplied by sex and the top ten principal components derived from the UK Biobank.

### Post GWAS Annotation and Analysis

The annotation of SNPs passing the GWAS significance threshold of 5e-8 and suggestive SNPs (p value 1e-6) was performed using FUMA version 1.4.0^87, 88^, ANNOVAR^89^, CADD v1.4^90^, RegulomeDB v1.1^91^, 15-core chromatin state^92^, eQTL and chromatin interaction mapping^93–96^ and expression GTEX database v8.

### Gene/Transcript Level Association Study

Gene-level association P values were computed using SNP-level summary statistics from GWAS output, using MAGMA v1.10^41^. Two rounds of analysis were performed, one in which gene-wide locations were used to annotate SNPs to genes, and the other in which transcript-wide locations were used to annotate SNPs with transcripts. SNPs were annotated to the gene level with the MAGMA annotation function and the reference data package with a 1 kb window upstream. Transcript locations were obtained using Canonical tables from the GENCODE v32 track in the UCSC Genome Browser database^97–99^ and were converted to Hg19 coordinates by LiftOver. A Bonferroni correction was applied in order to correct for multiple testing (considering 20K tests for gene level and 40K for transcript-based analysis.).

### Pre-Ranked Gene Set Enrichment Analysis

DGBI associated genes from the gene-based association testing method (Table S4) were sorted in descending order by the absolute value of their DGBI association z-score. Then, pre-ranked gene set enrichment analysis for the MSigDB Ontology gene sets was performed using fgsea v1.16. One tailed (scoreType “pos”) normalized enrichment scores were calculated for gene sets containing a minimum of 15 or maximum of 500 genes in each DGBI pre-ranked gene list. Gene sets significantly enriched in each DGBI pre-ranked gene list were identified based on a p-value cutoff < 0.01.

### Curation of Published Single Cell Transcriptomic Datasets

Curation of the snRNA-seq datasets of primary adult human colon and hPSC-derived stage 1 ganglioid and 2D enteric neuron cultures was conducted as previously described by our group^22, 23^. The cells from the control ganglioid (not pp121-treated) and vehicle-treated 2D enteric neuron samples were used in this study. For the primary fetal human enteric neuron scRNA-seq dataset, the normalized .H5AD file was downloaded from https://www.gutcellatlas.org/ and converted into a Seurat object. Datasets were analyzed in R version 4.2.1 with Seurat version 4.3.0^100^. For the primary human GI datasets, only the cells labeled as enteric neurons, enteric glia, ICCs, myocytes, or epithelial were used in our analyses. For the hPSC-derived enteric neuron datasets, only the cells labeled as enteric neurons were used in our analyses. Unless otherwise specified, the adaptively-thresholded low rank approximation (ALRA) imputed gene expression was used to evaluate gene expression in this study^101^.

### DGBI Transcriptional Scoring of GI Cell Types and Enteric Neuron Subtypes

To identify the expression level of the DGBI associated genes and transcripts in GI cell types, the cells from each human single cell transcriptomic dataset were scored for their expression of DGBIs molecular features. The cells were scored based on their expression of genes identified through our SNP-based genome wide association testing (mapped genes), transcripts identified through our gene-based genome wide association testing (z-score ± 3.5), and genes curated from previously published GWAS studies (published) (Table S5). Individual lists of genes or transcripts were used for each DGBI category as well as a combined list of genes or transcripts for each list generation method (mapped genes, z-score ± 3.5, and published). DGBI transcriptional scoring was performed using the “AddModuleScore” function in R version 4.2.1.

### Calculating Cell Type Transcriptional Signatures

*GI cell types.* The differentially expressed genes between each of the GI cell types (enteric neurons, enteric glia, ICCs, myocytes, and epithelial) was calculated from the non-imputed gene counts with the “FindAllMarkers” function using the Wilcoxon Rank Sum test and only genes with a positive fold change were returned.

#### Enteric neuron subtypes

The neurochemical identification of neurons was performed independently for each neurotransmitter and neuropeptide in each dataset. For each neurotransmitter, a core set of genes were selected consisting of the rate-limiting synthesis enzyme(s), metabolism enzymes and transport proteins (Table S8). Cells were first scored for each neurotransmission associated gene set using the “AddModuleScore” function. A cell was then annotated as “x-ergic” if the cell’s expression of a rate limiting enzyme was greater than 0 and the cell’s module score for the corresponding gene set was greater than 0. A cell was annotated as “Other” if both criteria were not met. For each neuropeptide, cells were scored for the gene corresponding with each neuropeptide using the “AddModuleScore” function (Table S8). A cell was then annotated as the neuropeptide if the cell’s expression of a gene was greater than 0 or “Other” if less than 0. The differentially expressed genes between each neurochemical identity and the “Other” population was calculated sequentially from the non-imputed gene counts with the “FindAllMarkers” function using the Wilcoxon Rank Sum test and only genes with a positive fold change were returned.

### Re-Clustering Analysis of GI Cell Types by DGBI Transcriptional Scores

To identify the cell types with high expression of DGBI molecular features, cells in the primary human GI datasets were scored based on their expression of transcripts identified through our gene-based genome wide association testing using the “AddModuleScore” function in R version 4.2.1. The cell by DGBI transcriptional score metadata from each dataset was merged and converted into a Seurat object. The variable feature sets were scaled and centered. Principal Components Analysis (PCA) was run using default settings and Uniform Manifold Approximation and Projection (UMAP) dimensionality reduction was performed using the PCA reduction with four principal components. The shared nearest neighbors (SNN) graph was computed using default settings and cell clustering was performed using the default Louvain algorithm at 0.15 resolution. The number of principal components used for UMAP reduction and SNN calculation was determined by principal component standard deviation. The average transcriptional score of each DGBI category was then evaluated for each cluster. The overall prevalence of each cell type represented in each cluster was calculated by summing the total number of cells in each cluster and calculating the percentage of each cell type represented in the cluster from this sum total.

### Gene-Set Analysis

We performed gene set analysis (GSA) using MAGMA on sets of differentially expressed genes between cell clusters. The model was corrected for gene size, density, MAC, and GWAS sample size. Competitive two-sided GSA was performed and Beta and SE of the output were used to calculate gene set association z-scores for each phenotype.

### Calculating DGBI Association Scores for Enteric Neuron Subtypes

For each human single cell transcriptomic dataset, the average DGBI module score of each enteric neuron subtype was scaled for each DGBI category. To generate an overall DGBI association score for each enteric neuron subtype, the DGBI specific scores from each dataset were averaged via the min-max scaled z-scores from the Gene-Set analysis approach.

Then the distance matrix of the combined table of subtype’s module scores and their corresponding DE gene-sets association was calculated using scipy version 1.8.0 and the relationship between clusters and the combined DGBI association score Visualized using multidimensional scaling in sklearn version 1.0.2.

### Protein-Protein Interaction Network Analysis

First we filtered our list of DGBI associated genes, curated from our SNP and gene-based genome wide association testing as well as the genes identified from previously published GWAS studies (Table S6), by their expression in the relevant DGBI associated enteric neuron subtypes (Figure 4D). Per human single cell transcriptomic dataset, each gene in the DGBI gene list was preserved based on the associated neurochemical ID having > 0% of its cells expressing the gene. For each DGBI category, only genes conserved across all four datasets per neurochemical identity were included in the downstream analysis. The neurochemical identity filtered DGBI gene lists were evaluated for protein-protein network interactions using the Search Tool for the Retrieval of Interacting Genes (STRING) database version 11.5. The minimum required interaction score was set to 0.4, corresponding to medium confidence with data support from the following active interaction sources: textmining, experiments, databases, co-expression, neighborhood, gene fusion, and co-occurrence.

### UCSF De-Identified Clinical Data Analysis

Looking at the nodes in DGBI pathways (Figure 5A) we identified drugs targeting them using the IUPHAR/BPS Guide to PHARMACOLOGY (GtoPdb) database^102, 103^. A full list of the drugs is provided in Table S7. DeID Clinical Data at UCSF^34^ comprises about 5 million patients. By using the same case/control definition and exclusion criteria as the UKBB cohort, we defined cases and controls and conducted a multivariate logistic regression analysis in order to determine if drugs targeting nodes have an influence on DGBI rates. Statistical testing was performed using statsmodels version 0.13.2 in Python version 3.8.8.

